# Personalized Feature Statistics: Individual-Level Variant Inference under Genetic Ancestry Continuum

**DOI:** 10.64898/2026.04.28.26351879

**Authors:** Julie Fangran Wang, Rachel Yu, Jacob Edelson, Junyoung Park, Yann Le Guen, Xiaoxia Liu, Michael E. Belloy, Iuliana Ionita-Laza, Michael D. Greicius, Hua Tang, Zihuai He

## Abstract

Genome-wide association studies (GWAS) have successfully identified numerous genetic variants associated with complex diseases. However, the extent to which the effects of these variants vary across populations of diverse ancestries remains poorly understood. Furthermore, in these contexts genetic ancestry is treated as a categorical variable, thereby oversimplifying its continuous nature and the more nuanced ways in which it can influence genetic effects on disease. Here, we propose personalized feature statistics (**PFstatistics**), a statistical framework that quantifies the importance of genetic variants to a phenotype based on each individual’s ancestry background, and profiles heterogeneous genetic effects across the genetic ancestry continuum. We demonstrate the utility of this framework through both simulations and real data analysis using sequencing data from ancestrally diverse cohorts in the Alzheimer’s Disease Sequencing Project (ADSP). We show that Alzheimer’s Disease (AD) risk variants span a spectrum from ancestry-homogeneous to ancestry-dependent effects, and that PFstatistics characterizes this spectrum at individual resolution across the ancestry continuum. PFstatistics also provides individual-level variant selection with FDR controlled at a target level, yielding distinct selection sets that vary across individuals according to their ancestry background. While demonstrated in the context of genetic ancestry, the proposed method is broadly applicable to other heterogeneity features such as environmental factors, offering a robust tool for understanding complex genetic contributions across diverse populations.

## Introduction

Genome-wide association studies (GWAS) have identified many genetic variants associated with complex diseases. An important unresolved question in human genetics is the extent to which these genetic effects are shared across populations or vary with ancestry background. Across many complex traits, effect sizes appear to be substantially, but not perfectly, shared across ancestries, and the degree of heterogeneity can vary by trait, locus, and genomic context. At the same time, differences in marginal effects across populations do not necessarily imply differences in underlying causal effects, because ancestry-specific linkage disequilibrium (LD) can induce apparent effect heterogeneity even when causal effects are similar.^1–3^

To study such heterogeneity, most existing work focuses on population-level summaries of genetic effects. Common approaches include stratified GWAS and multi-ancestry meta-analysis methods such as MR-MEGA.^4^ These methods have been useful for identifying loci with evidence of heterogeneity and for quantifying cross-ancestry differences in effect sizes. However, these approaches operate at the level of group- or population-average effects, relying on discrete ancestry labels and producing a single estimate or test statistic per variant, thereby failing to capture the continuous nature of human genetic variation. This limitation has motivated a broader shift toward modeling ancestry more continuously, including through local ancestry, principal components (PCs), ancestry proportions, and related genetic-distance representations.^5,6^

To capture finer-scale population structure, prior work has modeled admixed populations as mixtures of multiple ancestral components, but these approaches still rely on discrete groupings.^7^ More recent efforts have focused on continuous representations for genetic ancestry in different contexts, including polygenic risk score (PRS) prediction. For example, iPGS+refit, was developed to improve polygenic prediction in admixed individuals by combining ancestry-shared effects with ancestry-aware refitting when heterogeneous associations are present.^8^ Another method, SPLENDID incorporated genotype-by-ancestry interactions within a penalized regression framework to improve polygenic risk prediction across diverse populations.^9^ More recently, DiscoDivas leveraged the genetic ancestry continuum to interpolate PRS, with particular benefit in admixed individuals.^10^ Together, these studies show that replacing discrete ancestry groupings with continuous ancestry representations can improve PRS portability and applicability across diverse cohorts. However, these methods are designed primarily for prediction but do not address a different inferential question that is central to genetic studies: which variants are important for a given individual, how that importance changes across a continuous ancestry background, and how such feature selection can be performed with formal control of false discoveries.

To address this gap, we propose personalized feature statistics (**PFstatistics**), a framework for individual-level variant inference and FDR-controlled selection that characterizes how genetic effects vary continuously across individuals with different ancestry backgrounds. Within PFstatistics, we integrate Lasso-penalized regression with genotype-ancestry interaction effects, where ancestry is represented as a continuous variable by ancestry PCs and estimated ancestry proportions. We further integrate model-X knockoffs to perform conditional variable selection with false discovery rate (FDR) control. The model-X knockoffs framework provides two key advantages: first, it enables valid inference when traditional regression fails in high-dimensional or nonlinear settings; second, it provides formal FDR control, ensuring that selected variants are statistically justified. Together, these components enable PFstatistics to (1) quantify ancestry-driven heterogeneity on a continuous scale, and (2) select variants associated with a trait at the individual level. Moreover, rather than relying on a fixed, population-level significance threshold, PFstatistics uses adaptive, individualized selection thresholds to perform variant selection.

We study Alzheimer’s disease (AD) as a motivating application because it represents a particularly strong setting for ancestry-dependent genetic effects. In contrast to traits for which common-variant effects are often highly concordant across ancestries on average, AD includes well-documented examples of biologically meaningful heterogeneity, most notably at the APOE locus. Belloy et al. provided a large-scale analysis showing that APOE-associated AD risk varies across age, sex, race and ethnicity, and population ancestry, highlighting that this association is modulated by background factors rather than being constant across populations.^11^ This heterogeneity is not confined to *APOE*. Lake et al. performed a multi-ancestry meta-analysis of known AD loci and found that a substantial fraction of established risk regions showed significant heterogeneity across ancestry groups.^12^ Non-European GWAS have further reinforced this picture, identifying ancestry-specific signals in African American and African ancestry cohorts.^13,14^ However, these studies relied on discrete ancestry stratification, effectively applying a hard threshold, and neither accounted for sample admixture nor modeled ancestry as a continuous variable. As a result, the extent and structure of ancestry-dependent heterogeneity across the full ancestry continuum remains poorly characterized. This makes AD an especially informative and practically important setting in which to study individualized variant importance across the ancestry continuum.

Using simulations and sequencing data from the Alzheimer’s Disease Sequencing Project (ADSP), we show that PFstatistics can recover both ancestry-homogeneous and ancestry-dependent signals, provide individualized variant selection with controlled FDR, and reveal that AD risk architecture contains both ancestry-homogeneous loci and loci whose importance varies continuously across the ancestry background. Beyond variant inference, we further demonstrate that PFstatistics can diagnose PRS portability: by decomposing variant importance across the ancestry continuum, it identifies which variants drive portability loss and which individuals are most affected, providing a principled variant-level and individual-level explanation for why EUR-trained PRS underperforms in non-EUR populations. While demonstrated here in the context of genetic ancestry, the proposed framework is broadly applicable to other continuous sources of heterogeneity.

## Material and methods

### Motivating example: the ancestry continuum in the Alzheimer’s Disease Sequencing Project Follow-Up Study 2.0 (ADSP FUS 2.0)

We use the ADSP Release 5 WGS dataset as a motivating example to illustrate the continuous nature of human genetic ancestry. The ADSP is an NIH initiative integrating whole-genome sequencing data with phenotypic information across diverse populations to identify genetic factors influencing AD risk,^15^ with the Follow-Up Study 2.0 (FUS 2.0) specifically designed to expand representation beyond the predominantly non-Hispanic white samples in large biobank-scale GWAS.^16^

In this study, we considered 51,665 individuals with 27% AD cases and 73% non-AD cases. Self-reported races contain 49% Non-Hispanic White, 29% Hispanic or Latino, 13% Non-Hispanic Black or African American and 9% others. We derived ancestry principal components and estimated ancestry proportions for all individuals using the SNPweights framework,^17^ projecting onto the 1000 Genomes reference panel^18^ to obtain continuous ancestry coordinates in five dimensions (European (EUR), African (AFR), East Asian (EAS), South Asian (SAS) and Native American/Hispanic (AMR)). **Figure 1a** illustrates the resulting ancestry composition of the cohort. The smooth transitions between samples illustrate the continuous nature of human genetic variation. We then apply a 75% ancestry proportion threshold to define predominant-ancestry groups. Individuals are assigned to the ancestry group for which their estimated proportion exceeds 75%, and those who do not meet this criterion for any single ancestry are classified as Admixed. Shown in **Figure 1b**, the substantial fraction of admixed individuals emphasizes the need for methods that model ancestry continuously to better capture genetic heterogeneity. These discrete labels are used exclusively for visualization and for defining evaluation strata when comparing PFstatistics against ancestry-stratified benchmark methods. They are never used as inputs to PFstatistics, which operates entirely on continuous ancestry features.

**Figure 1.**
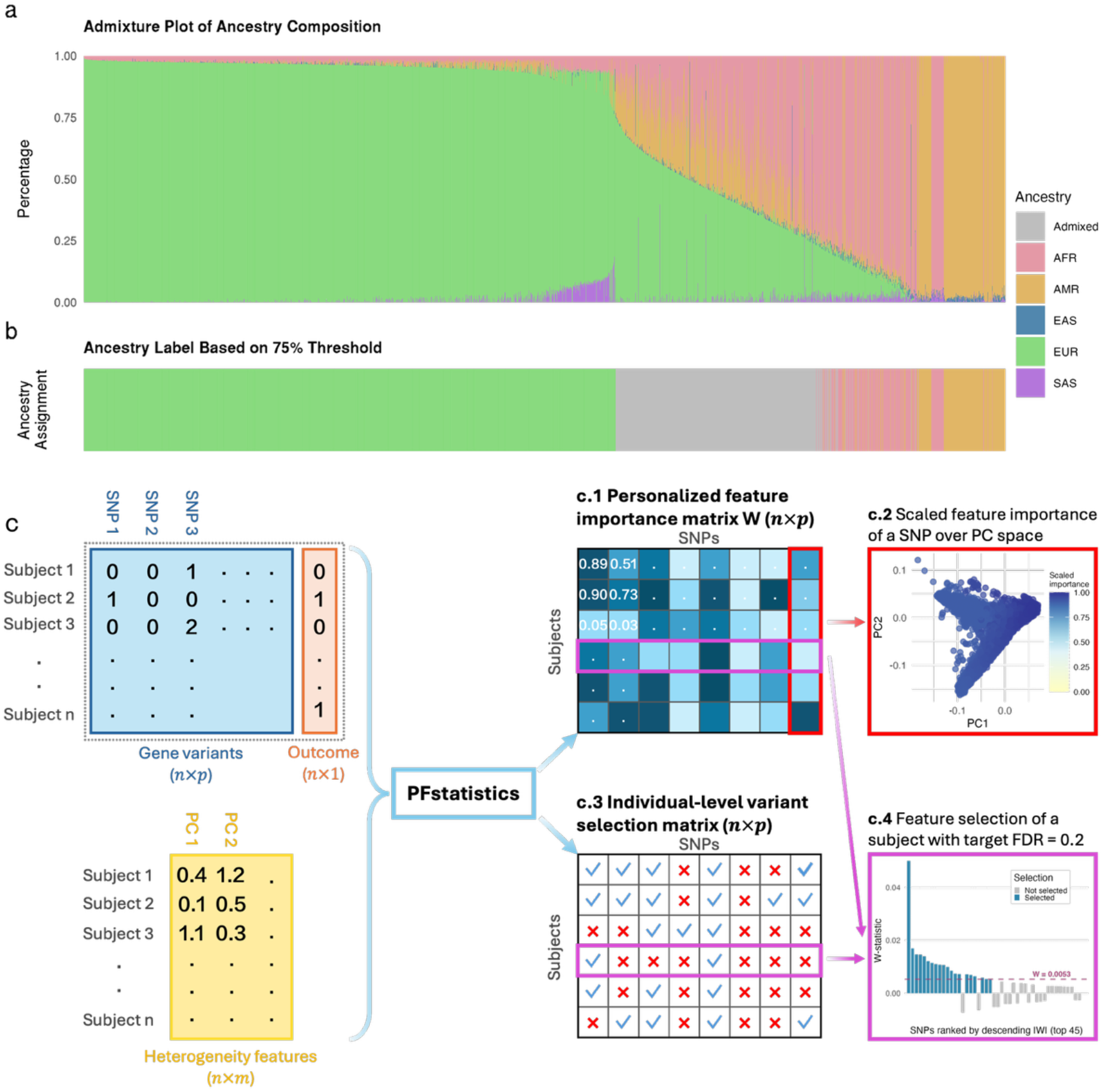
Overview of ancestry compositions and the PFstatistics framework. **a**. Admixture plot showing the continuous ancestry composition of all individuals across five groups (AFR, AMR, EAS, EUR and SAS). Each bar represents one subject, with color indicating proportional ancestry estimates. Subjects are sorted ascendingly by non-European ancestry. **b**. Discrete ancestry labels obtained by applying a 75% ancestry-proportion threshold. Individuals are classified as “Admixed” if no single ancestry exceeds 75%. The rest are assigned to their predominant ancestry group. **c**. Schematic of the PFstatistics workflow. Genotype data (*n* × *p*), outcome data (*n* × 1), and heterogeneity features such as principal components (*n* × *m*) are combined to produce a personalized feature-importance matrix (*n* × *p*) and an individual-level variant selection matrix (*n* × *p*), shown in **c.1** and **c.3**, respectively. **c.2** illustrates an example of per-SNP importance patterns across PC space, where each point corresponds to an individual, and color indicates the scaled feature importance. **c.4** presents an individual’s variant-selection result under a target FDR of 0.2; each bar presents a SNP’s feature importance, and the red dashed line presents the threshold for that individual.

### Candidate Variants

A central practical challenge in studying ancestry-dependent genetic effects is that large-scale GWAS, which have the statistical power to reliably discover and replicate risk loci, are predominantly conducted in European-ancestry populations, whereas the ancestrally diverse WGS cohorts better suited to characterizing heterogeneity across the ancestry continuum are typically smaller and underpowered for *de novo* genome-wide discovery. We address this by focusing on a candidate variant set that was previously identified through large-scale EUR-dominant GWAS and applying PFstatistics to evaluate their ancestry-dependent importance within the ADSP cohort. The candidate variants were further prioritized based on functional genomic evidence, including mapping AD risk variants in active myeloid enhancers to target genes, colocalization with expression quantitative trait loci (eQTL) in brain and immune tissues, and assessment of regulatory effects on molecular phenotypes in AD-relevant tissues and cell types.^19–21^ We retained variants common in the ADSP cohort (MAF > 5%) and applied hierarchical clustering to remove tightly linked variants, retaining one representative SNP from any cluster with pairwise correlation exceeding 0.9, yielding a final set of 107 candidate variants used in all subsequent simulation and real data analyses. This candidate set provides a realistic and interpretable feature space grounded in known AD biology and serves as a convenient pool of common variants with realistic LD structure for simulation purposes.

### PFstatistics: Personalized Feature Statistics for Genetic Studies

PFstatistics is a personalized variant testing and selection framework that addresses how a variant’s impact on a phenotype varies continuously along some heterogeneity features, such as genetic ancestry (**Figure 1c**).

#### Notation

Assume a study population of *n* independent subjects with *p* genetic variants. For the i-th subject, *Y*_*i*_ denotes a continuous phenotype variable; *G*_*i*,·_ ∈ *mG*_*i*1_, …, *G*_*ip*_) denotes the genotypes for the p variants; *Z*_*i*,·_ ∈ (*Z*_*i*1_, …, *Z*_*im*_) denotes the heterogeneity features, which can include principal components and estimated ancestry percentage as continuous representation of ancestry. Genotype *G*_*ij*_ typically represents the number of copies of minor allele and takes values of 0, 1, or 2.

#### Model heterogeneous SNP effects

To model the heterogeneous effects of variants, we include interaction effects between each genetic variant and heterogeneity feature in our linear model. Formally, the model writes as:

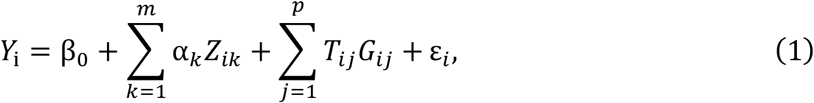

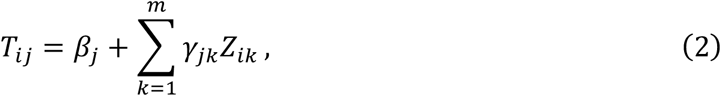

where *β*_0_ is intercept, *α*_*k*_ is the regression coefficient for the *k*-th heterogeneity feature, *T*_*ij*_ is the heterogeneity-aware coefficient for the j-th genotype, and *ε*_*i*_ is a random error. Note that *T*_*ij*_ is the gradient 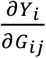 with respect to the *j*-th genetic variant for the *i*-th individual, which captures the sensitivity of the model output *Y*_*i*_ to perturbations in the input feature *G*_*ij*_ for a particular individual. *T*_*ij*_ breaks down into two parts: *β*_*j*_ denotes the main effect of j-th observed genotype and *γ*_*jk*_ is the coefficient for the interactions between the j-th genotype *G*_*ij*_ and the k-th heterogeneity feature *Z*_*ik*_. A larger magnitude of *T*_*ij*_ indicates greater influence of SNP *j* on individual *i*’s outcome. When there is no heterogeneous effect, i.e. *γ*_*jk*_ = 0 for all *j* and *k, T*_*ij*_ reduces to *β*_*j*_ which is a constant effect for all individuals. It is worth noting that additional covariates such as sex, age etc. can be included as in usual linear regression.

#### PFstatistics with penalized regression and model-X knockoffs

While the main objective is to evaluate *T*_*ij*_, classical linear regression methods break down when the number of predictors exceeds the sample size. This issue is exacerbated when interaction effects are included, due to the increased dimensionality of the feature space. In such high-dimensional settings, penalized regression methods such as the Lasso are frequently used to shrink the coefficients of irrelevant predictors toward zero, thereby performing regularization and aiding variable selection. However, while the Lasso can identify a sparse set of nonzero coefficients, it offers no guarantee of Type I error control. The selected features cannot be interpreted as statistically significant associations, and the false discovery rate remains uncontrolled.

PFstatistics overcomes these limitations by incorporating model-X knockoffs to perform variable selection with rigorous FDR control. The model-X knockoffs framework formulates the problem as a set of conditional independence tests with the null hypotheses

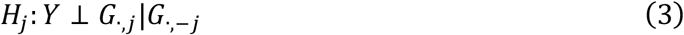

where *G*_·,−*j*_ = (*G*_·,1_, ⋯, *G*_·,*j*−1_, *G*_·,*j*+1_, ⋯, *G*_·,*p*_) contains all variants except j-th variant *G*_·,*j*_. We reject the null hypothesis *H*_*j*_ only if the phenotype is conditionally dependent on the j-th variant, suggesting *G*_·,*j*_ provides additional information to the phenotype once all other variants are accounted for.

To perform these conditional tests, model-X knockoffs first constructs knockoff replicas of genetic variants 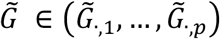 that are statistically indistinguishable from the originals but conditionally independent of the outcome variable given the original features. In this study, the knockoff variables are generated using the second-order model-X knockoffs construction introduced by Candes et al^22^, which matches the first two moments, i.e. mean and covariance, of the original variables and knockoffs.

To construct model-X knockoffs test statistics, PFstatistics uses Lasso-penalized regression as the learning model to test for association between the genetic variants and the phenotype. We consider an extended Lasso regression model that includes both the real and knockoff variants along with their interactions with heterogeneity features:

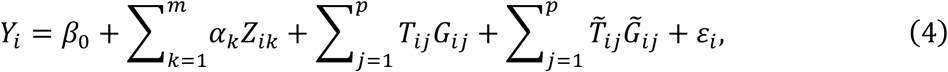

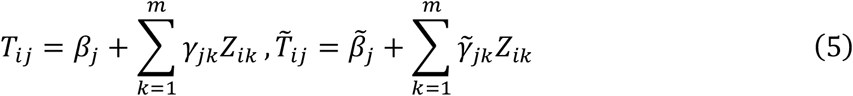

where 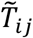 captures the effects of knockoff variables on the output. The extra parameters to be learned are 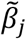, the coefficient for the main effect of the knockoff version of the j-th variant and 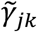, the coefficient for the interactions between the j-th knockoff and the k-th heterogeneity feature. For notation simplicity, we denote the linear predictor as

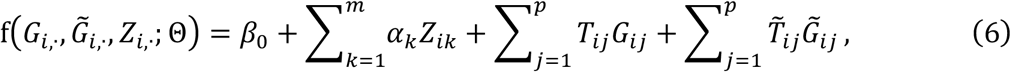

where 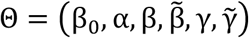 collects all parameters. Using this notation, the model is estimated by minimizing the Lasso objective:

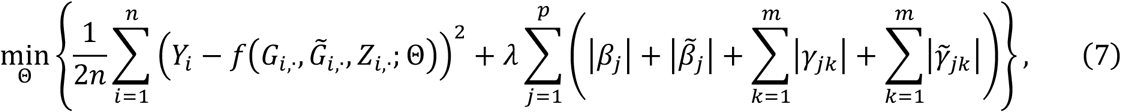

where *λ* is the regularization parameter. The ancestry main effects *α* are not penalized, allowing the model to adjust freely for population structure, while all genetic main and interaction effects (original and knockoff) are subject to *ℓ*_1_-penalization to induce sparsity. Model fitting was performed via a five-fold cross-validation using cv.glmnet, which selects the optimal regularization parameter *λ* to balance model sparsity and predictive performance.

In the extended model, the knockoff variants act as negative controls for the original variants. Under the null, the original and knockoff features are statistically indistinguishable in their relationship to *Y*. As for true variants, they are expected to exhibit higher influence on *Y* than their knockoff counterparts. We thus construct a knockoff test-statistic matrix, W-statistics (*W* ∈ *R*^*n*×*p*^), by contrasting the individual-level effects of the original variant with its knockoff:

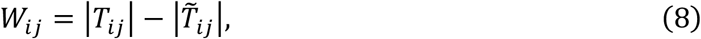

representing the *personalized feature importance matrix* (**Figure 1, panel c.1**) of a variant on individual level. The statistics depend on each subject’s unique profile and vary across samples (**Figure 1, panel c.2**). Intuitively, a large positive *W*_*ij*_ suggests that the original features have a stronger effect than the knockoffs and are more likely to be causal. Conversely, values near zero or negative imply that the feature may be spurious or redundant. To facilitate comparison across samples, we further define *scaled importance scores* across individuals as 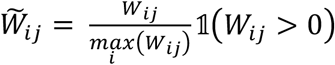, where 𝟙(⋅) is the indicator function. Only positive signals *W*_*ij*_ are considered because negative values suggest the *j*-th variant contributes no additional signal in the *i*-th individual.

Finally, the personalized feature selection by applying the knockoff filter^22^ separately to each subject, identifying putative causal features while controlling FDR for each individual. Specifically, for each sample *i*, the data-adaptive threshold is chosen as

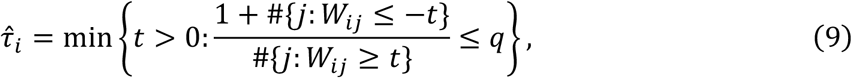

yielding a set of selected features 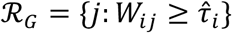 that controls the FDR at a target level *q*. We define a binary *individual-level variant selection* matrix *S* ∈ *R*^*n*×*p*^ that denotes selection results (**Figure 1, panel c.3**):

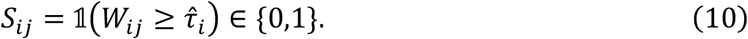

The selection sets depend on each subject’s distribution of feature importance, and the selection threshold varies across individuals (**Figure 1, panel c.4)**. We further illustrate this behavior in later sections through applications of PFstatistics to real-world data.

### Simulations

To assess the performance of PFstatistics, we conducted simulations designed to evaluate its ability to perform individualized feature selection with FDR control and to capture ancestry-dependent variation in variant importance along a continuous ancestry background.

We generated synthetic data based on a subset of the ADSP sequencing dataset to preserve realistic linkage disequilibrium (LD) structure. The pool of 107 candidate SNPs was used as input variants. To ensure that each causal variant contributed distinct signal and to avoid redundancy due to high correlation, we selected causal SNPs with low pairwise correlation. Specifically, we computed the absolute pairwise correlation matrix among candidate SNPs and performed single-linkage hierarchical clustering using a dissimilarity metric defined as 1 − |*r*|. A correlation threshold of 0.25 was used to define clusters, ensuring that SNPs selected from different clusters had pairwise correlations below this threshold. One representative SNP was randomly sampled from each cluster, and 30 SNPs were selected as causal variants.

Among these 30 causal SNPs, 10 were assigned homogeneous effects, while the remaining 20 were assigned ancestry-dependent effects. Specifically, 10 SNPs were designed to interact with European ancestry proportion and 10 with African ancestry proportion, allowing effect sizes to vary continuously across individuals.

We constructed a simulated cohort by randomly sampling 5,000 individuals from the ADSP dataset, including their genotypes *G*, principal components (PCs) and ancestry proportions, thereby preserving the underlying population structure. The outcome variable *Y*_*i*_ for individual *i* was generated using the following linear model:

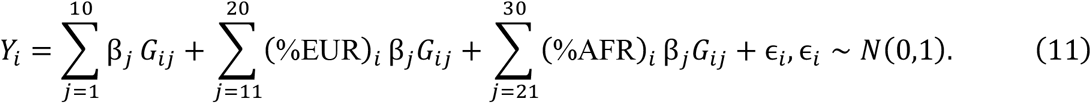

Here, the first 10 have homogenous effects across individuals (*β*_*j*_ = 0.1 or − 0.1), the next 10 interact with each individual’s European ancestry percentage (*β*_*j*_ = 0.13 or − 0.13) and the remaining 10 interact with African ancestry (*β*_*j*_ = 0.25 or − 0.25). This design creates a setting in which both ancestry-homogeneous and ancestry-dependent effects coexist.

These 30 causal variants are held fixed across all runs. The only source of variation between runs is the random error *ϵ*_*i*_. This consistency allows for comparisons of feature importance and selection decisions across simulation replicates. We performed 100 independent simulation runs. In each run, we simulated a new dataset {*G, Y*}, generated new knockoff variables 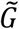 and applied our feature selection framework.

For each simulation run, we computed empirical power separately for each group of causal SNPs. For the 10 SNPs with homogenous effects, the empirical power for an individual is defined as the proportion of selected SNPs overlapping the homogeneous set of causal SNPs. This value varies across samples due to personalized selection. The empirical power for the ancestry-interacting groups was defined analogously. The false discovery proportion (FDP) for an individual is defined as the proportion of non-causal SNPs among all selected SNPs for that individual. Averaging the FDP across 100 simulation runs yields the empirical false discovery rate (FDR).

### Real data analysis

We demonstrate the effectiveness of PFstatistics in real-world data using AD as a modeling example due to its known ancestry-dependent genetic influences. We applied PFstatistics to the full ADSP Release 5 WGS dataset (*n* = 51,665) using the 107 candidate SNPs described above. We use the top four principal components and five estimated ancestry proportions as the heterogeneity features. The method yields personalized variant importance (W-statistics) and individual-level variant selection results. We also conduct marginal tests to verify the results.

## Results

### Simulations: Profiling continuous heterogeneous genetic effects

We conducted simulations to evaluate PFstatistics: whether the per-individual FDR is controlled at the nominal level, and whether the personalized W-statistics correctly reflect the structure of ancestry-dependent genetic effects. Simulations used genotype data from ADSP under a model with homogeneous and ancestry-dependent causal variants and results are averaged over 100 independent runs (see **Methods**).

PFstatistics controls the per-individual FDR at the nominal level across all individuals. The empirical FDR remains tightly controlled under the target level of 0.2 regardless of ancestry background and under ancestry-interacting causal effects (**Figure 2a**), supporting the validity of the personalized selection framework under structured population settings.

**Figure 2.**
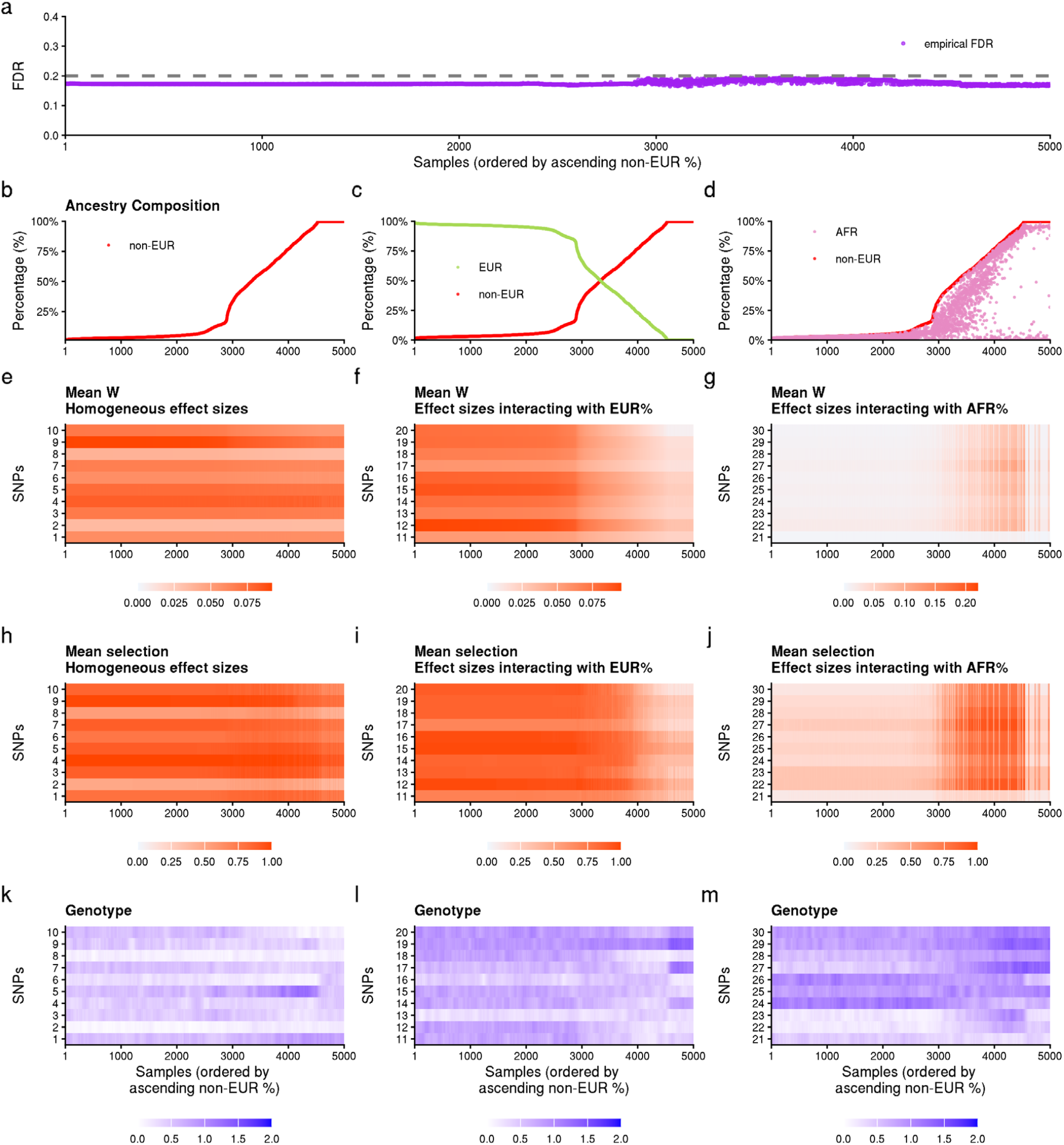
Simulation results. **a**. The empirical FDR across the samples with the dashed line denoting the target FDR controlled at 0.2. **b-d**. Sample ancestry composition distribution. All samples are ordered by increasing non-European ancestry proportion, defined as (1 − %EUR). **b** shows non-EUR ancestry composition, **c** contrasts EUR and non-EUR ancestry proportions and **d** contrast AFR and non-EUR ancestry proportions. **e-g**. Mean W-statistics of 100 simulation runs across SNPs that in simulated model, have homogeneous effect sizes, effect sizes interacting with European (EUR) ancestry proportion and effect sizes interacting with African (AFR) ancestry proportion, respectively. Within each panel, SNPs are numbered from small to large according to ascending coefficients of variation (standard deviation divided by mean) of their mean W-statistics. **h-j**. Mean selection results of 100 simulation replications across SNPs included in panels e-g, respectively. **k-m**. Genotype (minor allele) heatmaps that for SNPs included in panels e-g, respectively. Color intensity represents the average genotype within a sliding window of 100 individuals with similar non-European ancestry proportion.

The personalized W-statistics recover the true effect structure of each SNP class (**Figure 2e–j**). SNPs with homogeneous effects show consistent W-statistics and selection probabilities across individuals, regardless of ancestry compositions (**Figure 2e, 2h**). SNPs whose effects interact with European ancestry show W-statistics that increase smoothly with EUR ancestry proportion and decrease toward zero in individuals with predominantly non-European backgrounds (**Figure 2f, 2i**). The symmetric pattern holds for AFR-interacting SNPs (**Figure 2g, 2j**). Importantly, these are smooth, continuous gradients across individuals and reflect unique genetic effects in individuals.

The power to detect ancestry-interacting effects depends on allele frequency and effect size. Rare variants with low minor allele frequency, particularly within a given ancestry stratum, may show attenuated W-statistics despite having true interacting effects, because insufficient genotype variation limits the model’s ability to estimate ancestry-specific contributions. This is illustrated in the MAF differences in ancestry-stratified groups in **Figures S1–S3**. When allele frequencies are sufficiently high across ancestry groups, PFstatistics robustly recovers ancestry-dependent effect gradients even when those frequencies differ across groups (**Figure 2f**, SNP 13).

We further investigated how PFstatistics handles LD when tagging SNPs are present in the dataset. We started with defining tagging SNPs as the top 15 non-causal variants with the largest absolute correlation (correlation range: 0.58-0.89) with any of the 30 causal SNPs. Two scenarios were examined: (1) training data contained both causal and tagging SNPs, and (2) training data contained only tagging SNPs. **Figure S4** summarizes the average W-statistics and selection decisions across 100 replicates. When causal SNPs were included in training, our method correctly prioritized the causal variants and did not erroneously assign effects to tagging SNPs. By contrast, when causal SNPs were excluded from the training set, tagging SNPs naturally tended to be selected, and their inferred heterogeneous patterns closely mirrored those of the corresponding causal SNPs (**Figure S5**). As a further investigation, we simulated phenotypes using the same 30 causal SNPs but restricted to all-homogeneous, all EUR-interacting or all AFR-interacting causal effects. In this design, the 30 causal SNPs were again excluded from training, leaving 77 non-causal SNPs for selection. Results (**Figure S6–S8**) show that tagging SNPs selected by our method exhibited effect patterns consistent with their corresponding causal variants. Together, these analyses demonstrate that our method is robust in capturing underlying genetic signals, whether causal SNPs are directly observed or only represented through tagging SNPs.

### Profiling ancestry-heterogeneous AD risk variants in ADSP data

A key output of PFstatistics is a continuous measure of variant importance that delineates, at the individual level, which variants contribute to AD risk and how these effects vary as a function of ancestry. We applied PFstatistics to the full ADSP cohort (*n* = 51,665) using 107 candidate AD variants and ancestry PCs and estimated ancestry proportions as heterogeneity features. **Figure 3a** shows the genetic ancestry continuum of the ADSP cohort on the space of first two PCs. Distinct clusters appear on the extremes of the PC space for AFR, EUR, AMR, EAS, and SAS individuals, while admixed individuals are concentrated in the central region.

**Figure 3.**
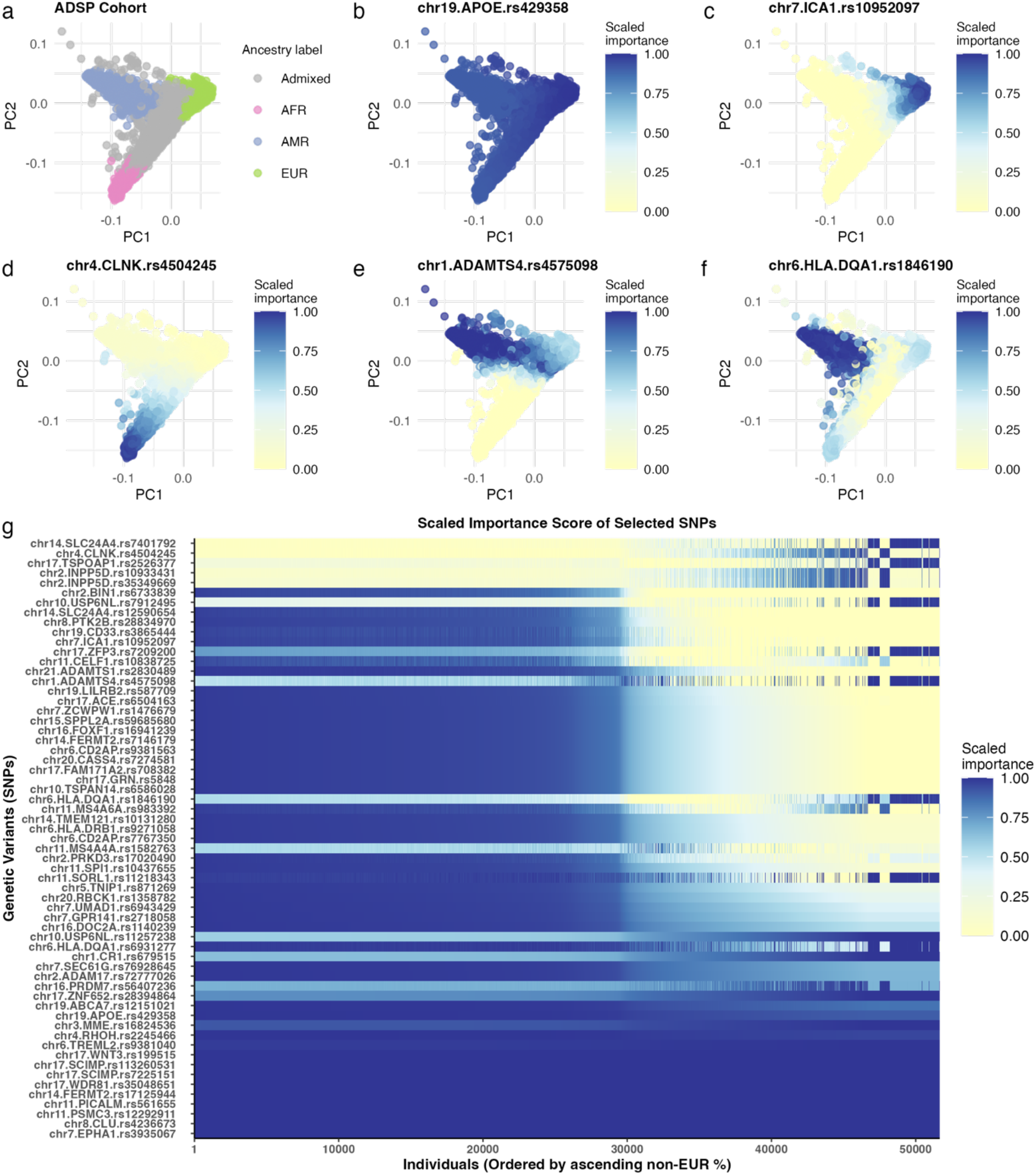
Profile continuous heterogeneous risk effects of ADSP cohort. **a**. PCA plot (PC1 and PC2) of the ADSP cohort, with individuals color-coded by ancestry label defined by the dominant ancestry composition (>75% cutoff). The plot illustrates genetic diversity and ancestry continuum within the cohort. **b–f**. Scaled importance scores for selected SNPs projected onto the same PC1–PC2 space. *APOE* and *ICA1* exhibit the strongest importance within the EUR cluster. *CLNK* shows elevated importance in the AFR cluster. *ADAMTS4* and *HLA-DQA1* display stronger importance toward the AMR cluster. **g**. The heatmap shows scaled importance scores for SNPs selected in at least one individual in the cohort, with patterns that gradually change across individuals ordered by their non-European ancestry proportions. SNPs are arranged top to bottom by descending heterogeneity of scaled importance, measured by the coefficient of variation, defined as standard deviation divided by mean.

To illustrate how variant importance varies across individuals, we projected the scaled importance scores 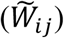 for five example AD loci onto the same PC space (**Figure 3b-f**). The resulting maps reveal qualitatively distinct patterns across different SNPs. For example, *APOE* SNP rs429358 which defines the *APOE* ε4 allele has been extensively studied for its strong association with increased AD risk in European populations and its effect can be weaker across other ancestries.^23^ PFstatistics recapitulates this pattern with more detailed ancestry contribution: individuals towards the European ancestry cluster exhibit the highest importance scores for this SNP, with signal gradually tapering off among admixed and non-European populations (**Figure 3b**). *CLNK* shows elevated importance in the AFR cluster (**Figure 3d**), while *ADAMTS4* and *HLA-DQA1* show stronger importance toward the AMR cluster (**Figure 3e, 3f**), illustrating that ancestry-dependent effects differ in their directionality across loci.

Of the 107 candidate variants, 61 SNPs were selected in at least one individual at FDR = 0.2 across the cohort, with an average of 42.6 SNPs selected per individual (range: 6-60). We measured a SNP’s heterogeneity using the coefficient of variation (CV) of its scaled importance, defined as the standard deviation divided by the mean. Higher CV reflects stronger ancestry-dependent heterogeneity, while lower CV reflects more uniform importance across the continuum. **Figure 3g** arranges these 61 SNPs by descending CV across individuals ranked by non-European ancestry proportion (non-EUR%). SNPs at the top show importance scores that vary strongly and systematically with ancestry background. SNPs toward the bottom display largely uniform importance across individuals regardless of ancestry, consistent with ancestry-homogeneous effects. Taken together, these results show that AD risk architecture contains both ancestry-heterogeneous and ancestry-homogeneous loci, and that PFstatistics can characterize where individual variants fall along this spectrum at individual resolution.

### Individual-level feature selection and personalized risk stratification

PFstatistics enables individual-level variant selection with explicit FDR control. **Figure 4** shows the W-statistic profiles for four representative individuals spanning distinct ancestry backgrounds (EUR, AFR, AMR, and Admixed). Across these individuals, PFstatistics yields distinct distributions of personalized feature importance (W-statistic), prioritizing different leading AD risk variants. Applying the data-adaptive W-statistic threshold separately for each individual at a target FDR of 0.2 results in distinct selection sets of risk variants. This demonstrates that PFstatistics can prioritize individual-level genetic risk factors with formal statistical control.

**Figure 4.**
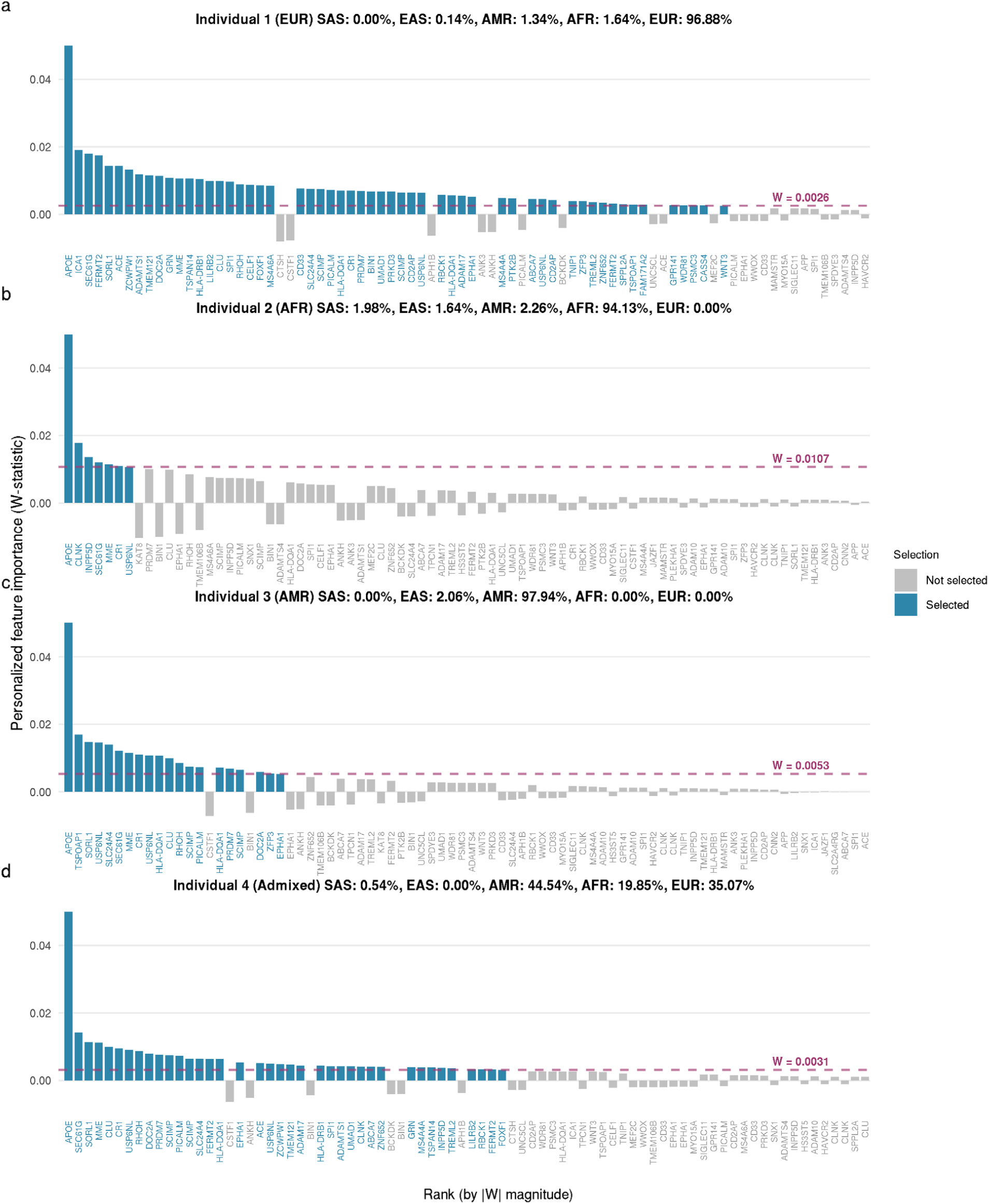
Personalized feature importance (W-statistic) profiles vary across individuals. **a–d**. show ranked personalized feature importance (W-statistics) for candidate SNPs from four representative individuals belonging to major ancestry groups: EUR, AFR, AMR and Admixed. The estimated ancestry proportions for each individual are shown in the panel titles. Each bar represents the W-statistic of a SNP, ordered by decreasing absolute magnitude (|W|). For better visualization, only the 80 SNPs with the largest |W| values are shown. The importance values of APOE SNP are capped at 0.05 to facilitate visualization. Blue bars denote SNPs selected above the individual-specific variant selection threshold (horizontal dashed line), whereas gray bars indicate unselected SNPs. Variation in selection thresholds across individuals reflects both ancestry-dependent differences in genetic architecture and stochastic variability inherent to the knockoff procedure.

Smaller selection sets in some individuals do not necessarily indicate fewer underlying risk variants but reflect the degree to which the available data can support inference for that individual’s genetic background. Individuals with well-represented ancestries in the training cohort (e.g., EUR) exhibit larger selection sets. In contrast, individuals from underrepresented groups (e.g., AFR and AMR) show more conservative selection sets, reflecting limited power to resolve ancestry-specific signals rather than a reduced burden of causal variants. This distinction highlights a key advantage of PFstatistics: it explicitly encodes uncertainty and avoids overconfident inference in settings where the data are less informative. As sequencing cohorts expand to include more diverse individuals, we expect selection sets to grow and variant rankings to stabilize for non-European individuals. A comprehensive view of individual-level selection patterns across the full cohort is provided in **Figure S9**.

### Diagnosing PRS portability

A key limitation of existing PRS is that their reduced predictive accuracy in non-European individuals is typically characterized as an aggregate performance gap at the population level, but the variant-level determinants of this portability loss remain poorly understood. PFstatistics addresses this directly: by quantifying each variant’s importance as a continuous function of ancestry, it identifies which variants drive ancestry-specific associations and which are robust cross-ancestry signals. This variant-level decomposition provides a principled explanation for PRS portability gaps and a framework for identifying which individuals are most affected (**Figure 5**).

**Figure 5.**
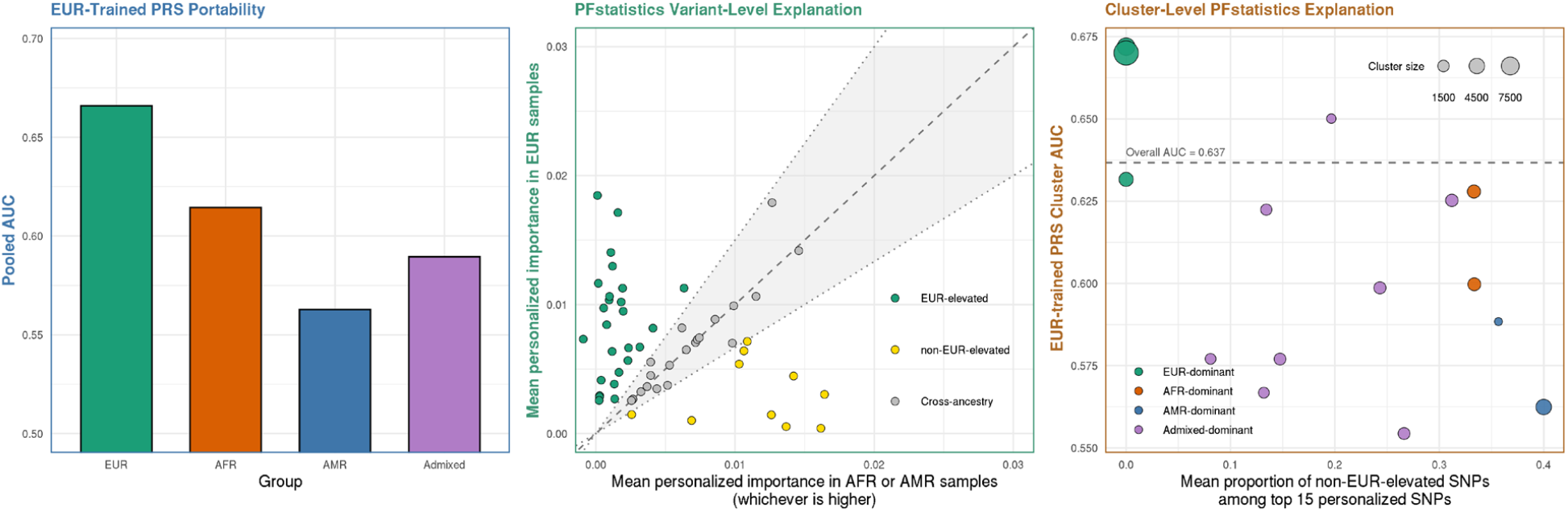
PFstatistics provides a variant-level and individual-level explanation for EUR-trained PRS portability loss. **a**. Performance of the EUR-trained PRS across ancestry groups, measured as pooled AUC in EUR, AFR, AMR, and Admixed individuals. The EUR-trained model performs best in EUR samples and shows reduced discrimination in non-EUR groups, demonstrating a portability gap. **b**. PFstatistics variant-level explanation of PRS portability. Each point represents one selected SNP, plotted by its mean personalized importance in EUR individuals versus the larger of its mean personalized importance in AFR or AMR individuals. *APOE* is excluded for a zoom-in visualization. SNPs are colored according to a threshold-based classification: a SNP is labeled EUR-elevated if its mean importance in EUR is greater than 1.5 times the larger of its mean importance in AFR or AMR; it is labeled non-EUR-elevated if the larger of its AFR or AMR mean importance is greater than 1.5 times its EUR mean importance; otherwise it is labeled Cross-ancestry. The shaded band marks the cross-ancestry region. **c**. PFstatistics individual-level decomposition of PRS portability. Individuals were clustered using k-means method on their personalized PFstatistics importance (W-statistics) profiles, and each point represents one cluster. The y axis shows cluster-level AUC of the EUR-trained PRS. The x axis shows the mean proportion, across individuals in that cluster, of top 15 personalized SNPs that are classified as non-EUR-elevated under the Panel B definition. Point color denotes the dominant ancestry composition of the cluster. Point size is proportional to cluster size. Dashed line denotes the AUC evaluated in all out-of-fold samples.

To illustrate this, we analyzed an Alzheimer disease PRS trained in European-ancestry samples and evaluated its performance across EUR, AFR, AMR, and Admixed individuals. As expected, the EUR-trained PRS showed the highest discrimination in EUR samples, with reduced performance in the non-EUR groups, confirming a clear portability gap (**Figure 5a**). We then leveraged the W-statistic matrix to dissect this portability pattern at the variant level. For each selected variant, we compared its mean personalized importance in EUR individuals against the larger of its mean importance in AFR or AMR individuals. Using a 1.5-fold importance ratio as the classification threshold, this analysis revealed a mixture of EUR-elevated, non-EUR-elevated, and cross-ancestry signals, rather than a uniform attenuation of predictive effects across ancestries (**Figure 5b**).

At the individual level, we clustered subjects by their PFstatistics importance profiles and examined the relationship between cluster-level PRS discrimination and the proportion of non-EUR-elevated variants among each individual’s top-ranked personalized signals (**Figure 5c**). Each cluster is labeled by the ancestry group that predominates among its members. Clusters with a higher burden of non-EUR-elevated variants tended to exhibit lower EUR-trained PRS discrimination, consistent with the hypothesis that portability loss is partly driven by variant-level ancestry dependence. Some AFR-, AMR-, and Admixed-dominant clusters retained moderate PRS discrimination despite carrying a relatively high proportion of non-EUR-elevated variants, indicating that PRS portability loss is not determined solely by ancestry label or by the presence of ancestry-elevated variants. Instead, these results suggest that PRS portability reflects the combined effects of cross-ancestry and ancestry-elevated variant importance within an individual, and that PFstatistics provides a variant-level framework for understanding where and why portability degrades and an individual-level characterization of which subjects are most susceptible to that degradation, across the ancestry continuum.

### Connections between PFstatistics and conventional marginal association tests

We connect PFstatistics with marginal association tests commonly used in GWAS by comparing their estimated SNP importance scores across stratified ancestry groups. Specifically, for each SNP and each ancestry stratum (Overall, EUR, AFR, AMR, and Admixed), we fit Gaussian marginal regression models within ancestry-stratified samples, adjusting for sex, ancestry principal components, and estimated ancestry proportions, and define the resulting regression coefficients as marginal importance scores.

Because PFstatistics targets conditional genetic effects rather than marginal associations, a direct comparison requires an intermediate representation. To bridge marginal analyses and PFstatistics, we fit a Lasso interaction model on the full sample that includes SNP-by-ancestry covariate interaction terms (**Eq. 1**) and compute individual-level conditional importance estimates as defined in **Eq. 2**. We then obtain ancestry-specific conditional importance scores by averaging these individual-level estimates within each ancestry group. Comparing the ancestry-stratified conditional importance scores with the corresponding marginal importance scores reveals strong overall agreement across SNPs, as reflected by high correlations (**Figure 6a**), indicating that conditional modeling preserves the dominant marginal signals while accounting for ancestry-dependent heterogeneity.

**Figure 6.**
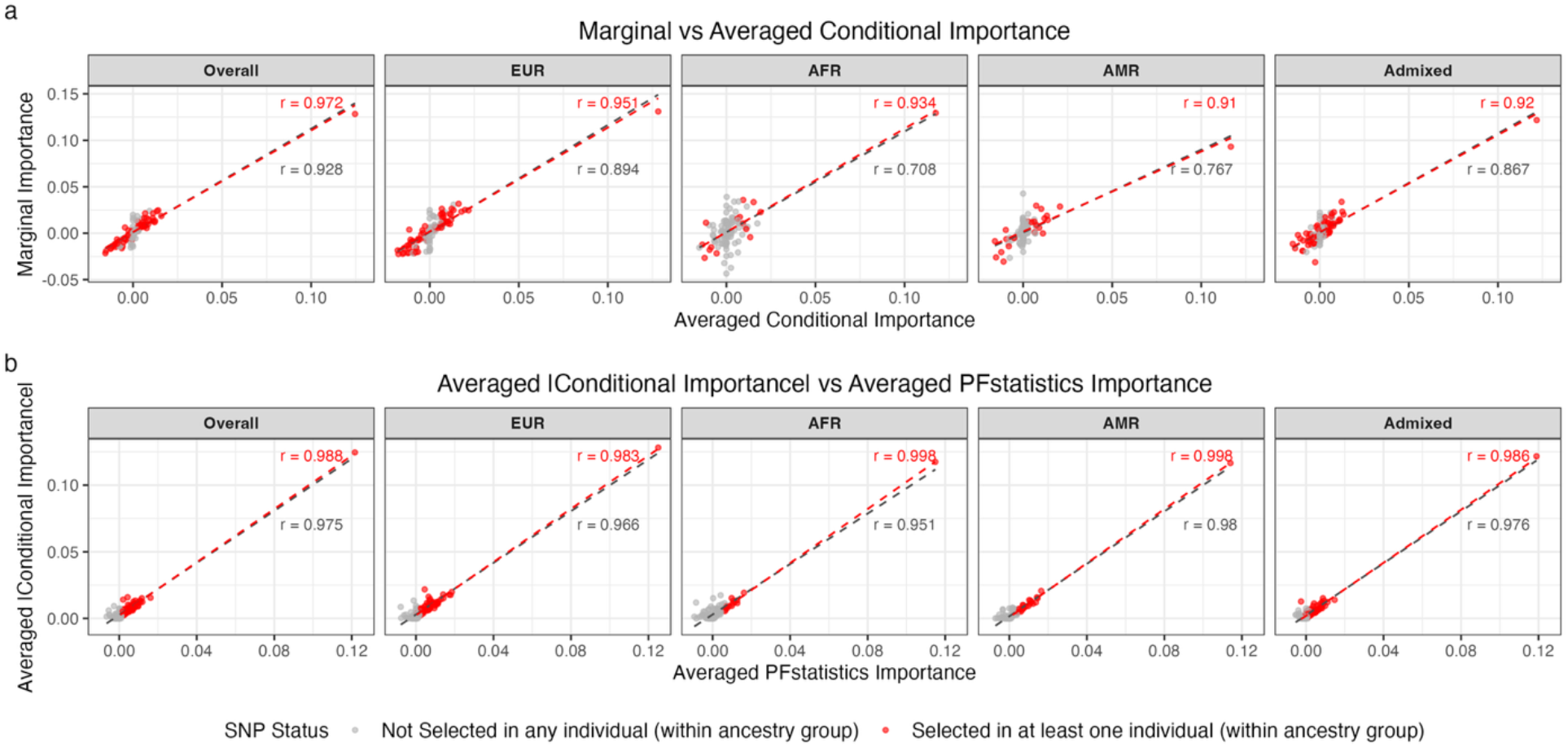
Comparison between marginal, conditional, and PFstatistics importance scores across ancestry groups. **a**. Scatterplots comparing marginal importance scores from ancestry-stratified Gaussian regression models with conditional importance scores derived from the Lasso interaction model, shown separately for the Overall sample and for each ancestry group (EUR, AFR, AMR, and Admixed). **b**. Scatterplots comparing in stratified groups the averaged absolute conditional importance scores with ancestry-averaged knockoff importance scores (W-statistics) produced by PFstatistics for the same SNP set. In both panels, each point represents a SNP. Red points indicate SNPs selected in at least one individual by PFstatistics within the corresponding ancestry group, whereas gray points represent SNPs not selected by PFstatistics in any individuals in that group. Red dashed lines represent the regression fitted to the selected SNPs (red points) within each ancestry group, while the gray dashed line represents the regression fitted to all SNPs. Pearson correlation coefficients (r) are reported for selected SNPs (red texts) and for all SNPs (grey texts), quantifying agreement between importance measures within each ancestry stratum.

Since the PFstatistics model (**Eq. 4**) is a direct extension of the conditional importance framework (**Eq. 2**), we next compare PFstatistics personalized importance scores (W-statistics) with conditional importance scores. We analyze the importance scores produced by PFstatistics on the same SNP set and compute ancestry-specific PFstatistics importance scores by averaging W-statistics within each ancestry stratum. Because the W-statistic encodes effect magnitude without direction, we compare PFstatistics importance scores with the averaged absolute conditional importance scores within each ancestry group. Across ancestry groups, the knockoff-based importance scores show strong concordance with the conditional importance scores (**Figure 6b**), demonstrating that PFstatistics recovers the same underlying conditional genetic signals while enabling controlled feature selection. As expected, knockoff-based inference introduces additional variability into importance score estimation as a trade-off for false discovery rate control. In **Figure S10**, we further present the same analyses with the *APOE* SNP excluded, which appears as the upper-right point and dominates the regression in each panel. The consistent agreement between marginal, conditional, and PFstatistics importance scores described above remains preserved.

## Discussion

We presented PFstatistics, a framework for individual-level variant inference and selection with FDR control that enables characterization of how variant importance varies across a continuous ancestry background. In contrast to existing methods that estimate population-level effect sizes per ancestry group, PFstatistics assigns each individual their own variant importance scores that reflect their specific ancestry composition and applies a per-individual adaptive threshold to make variant selection decisions with controlled false discovery rates. Using simulations and whole-genome sequencing data of candidate variants from ADSP, we show that PFstatistics recovers both ancestry-homogeneous and ancestry-dependent genetic signals across an ancestry continuum, maintains FDR control under structured population settings, and reveals that AD risk architecture contains both ancestry-heterogeneous loci, such as *APOE*, and ancestry-homogeneous loci whose importance is stable across the ancestry continuum.

Simulations further reveal that the power to detect ancestry-interacting effects depends on allele frequency. When allele frequencies are low within a given ancestry stratum, there is insufficient genotype variation to reliably characterize ancestry-dependent effects, producing signals that appear more homogeneous than they are. This limitation is expected to improve with larger sample sizes, where more abundant signal across the ancestry continuum enables better characterization of ancestry-interacting effects.

Beyond variant inference, PFstatistics offers a principled framework for diagnosing why PRS portability degrades across populations. A persistent challenge in polygenic prediction is that EUR-trained PRS underperforms in non-EUR individuals, yet the variant-level and individual-level determinants of this gap are rarely characterized. PFstatistics addresses this by decomposing variant importance across the ancestry continuum, revealing that portability loss is affected by a heterogeneous mixture of EUR-elevated, non-EUR-elevated, and cross-ancestry signals in different individuals. As a proof-of-concept extension, we evaluated PFstatistics-based PRS construction in simulations and in the ADSP cohort (**Supplemental Note**). While PFstatistics PRS showed advantages in simulations, differences in predictive accuracy were modest in real data, reflecting the inferential rather than predictive focus of the framework. PFstatistics is designed for individual-level variant inference and selection, not for maximizing prediction. Developing ancestry-adaptive PRS that more fully leverages the personalized importance estimates from PFstatistics remains a promising direction for future work.

As with other knockoff-based methods, the stochastic nature of knockoff generation can introduce variability in selected variant sets across repeated runs. This variability is more pronounced for variants with weaker effects, including those with low minor allele frequency in some ancestry stratum, while variants with stronger effects tend to be selected more consistently. Rather than representing a flaw specific to PFstatistics, this behavior highlights broader challenges in high-dimensional inference under correlation. Several extensions may improve stability and reproducibility, including refining knockoff generation procedures, exploring alternative regularization strategies, and employing multiple knockoffs to stabilize inference^24–26^. Ensemble approaches that aggregate results across multiple runs may also provide more robust assessments of feature importance. These directions represent promising avenues for future methodological development.

The present study intentionally restricts analysis to a curated set of candidate variants from European-dominant GWAS, reflecting a fundamental asymmetry in current multi-ancestry genomic resources: large-scale GWAS provide the statistical power needed for robust variant discovery but are predominantly EUR-ancestry, while ancestrally diverse WGS cohorts offer the population heterogeneity needed for characterizing ancestry-dependent effects but are currently too small for genome-wide inference. Analyzing established candidate variants in a diverse WGS cohort is therefore a practical strategy that allows PFstatistics to operate on a biologically motivated and statistically tractable variant set. As diverse sequencing cohorts grow in scale, extending PFstatistics to genome-wide analyses becomes increasingly feasible. Lasso-based penalized regression has already been demonstrated at genome-wide scale when sample sizes are sufficiently large,^9,27,28^ and the PFstatistics framework is directly compatible with such extensions. Applying PFstatistics genome-wide in well-powered diverse cohorts will be an important next step toward a more comprehensive characterization of ancestry-dependent variant importance across the full genetic architecture of AD and other complex traits.

From a computational perspective, PFstatistics demonstrates favorable performance for moderate-scale individual-level analyses. In our application, the evaluation of 51,665 individuals and 107 genetic variants with nine confounding features completed in approximately 354 seconds using base R in a single-threaded environment on an Apple M1 Pro chip (16 GB RAM). This runtime includes approximately 4.4 seconds for knockoff generation and 350 seconds for deriving importance matrices (∼258 seconds for fitting Lasso model and ∼80 seconds for applying knockoff filter). While efficient for datasets of this scale, extending PFstatistics to genome-wide analyses involving millions of variants presents additional computational challenges. Future work could leverage parallelization, distributed computing, sparse matrix representations, and more efficient knockoff generation algorithms to improve scalability.

While this study focused on modeling genetic ancestry as the primary heterogeneity feature, PFstatistics is flexible and can be extended to model other sources of heterogeneity in complex traits. Continuous environmental variables, socioeconomic factors, and other contextual covariates can be naturally integrated into the framework, which allows researchers to study their interactions with genetic variants. This generality broadens the applicability of PFstatistics beyond AD, positioning it as a versatile tool for investigating heterogeneous factors across a wide range of phenotypes.

## Data Availability

All data are available in the Alzheimer's Disease Sequencing Project (ADSP).

## Acknowledgments

Z.H. was supported by NIH/NIA award AG089509, AG066206 and AG066515.

## Data and code Availability

The data supporting the findings of this study are available from the Alzheimer’s Disease Sequencing Project (ADSP) and affiliated databases.

We have implemented PFstatistics (Personalized Feature Statistics for Genetic Studies) in a computationally efficient R package that can be applied generally to the analysis of other whole-genome sequencing studies. The package can be accessed at https://github.com/juliefrwang/PFStatistics.

## Supplemental Information

### Supplemental Figures

**Figure S1.**
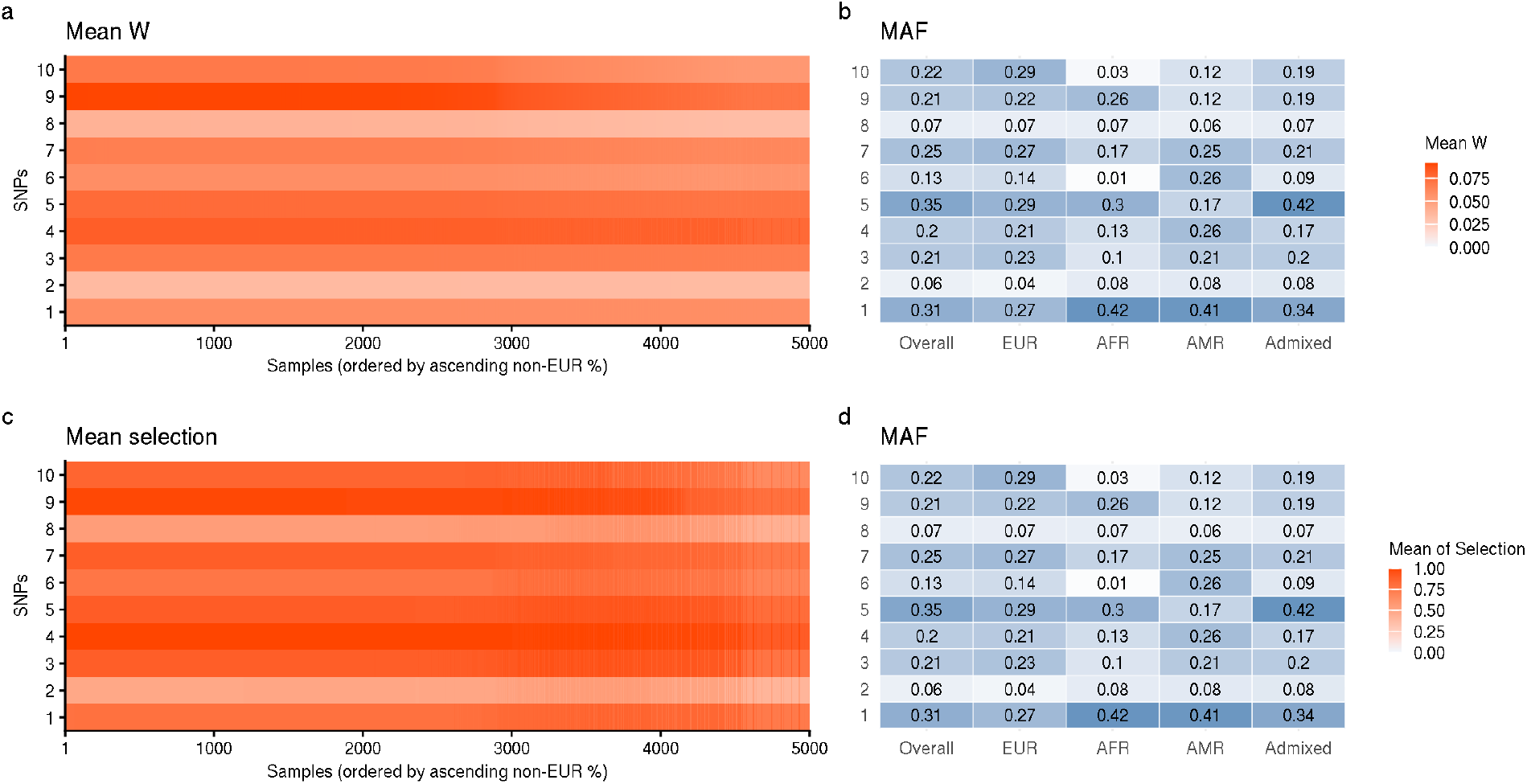
Minor allele frequencies (MAF) of homogenous causal variants across stratified groups and the full samples. In panels a-d, the Y-axis corresponds to the 10 homogeneous causal variants used in the simulation settings described in Section 2.3. Panels **a** and **c** are the results shown in **Figure 2**, panels d and g. Panels **b** and **d** display the minor allele frequencies (MAF) of these causal SNPs across different ancestry groups and in the overall sample. Groups labeled in X-axis of b and d are defined by the estimated ancestry proportion greater than 75%; individuals not meeting this threshold are categorized as admixed.

**Figure S2.**
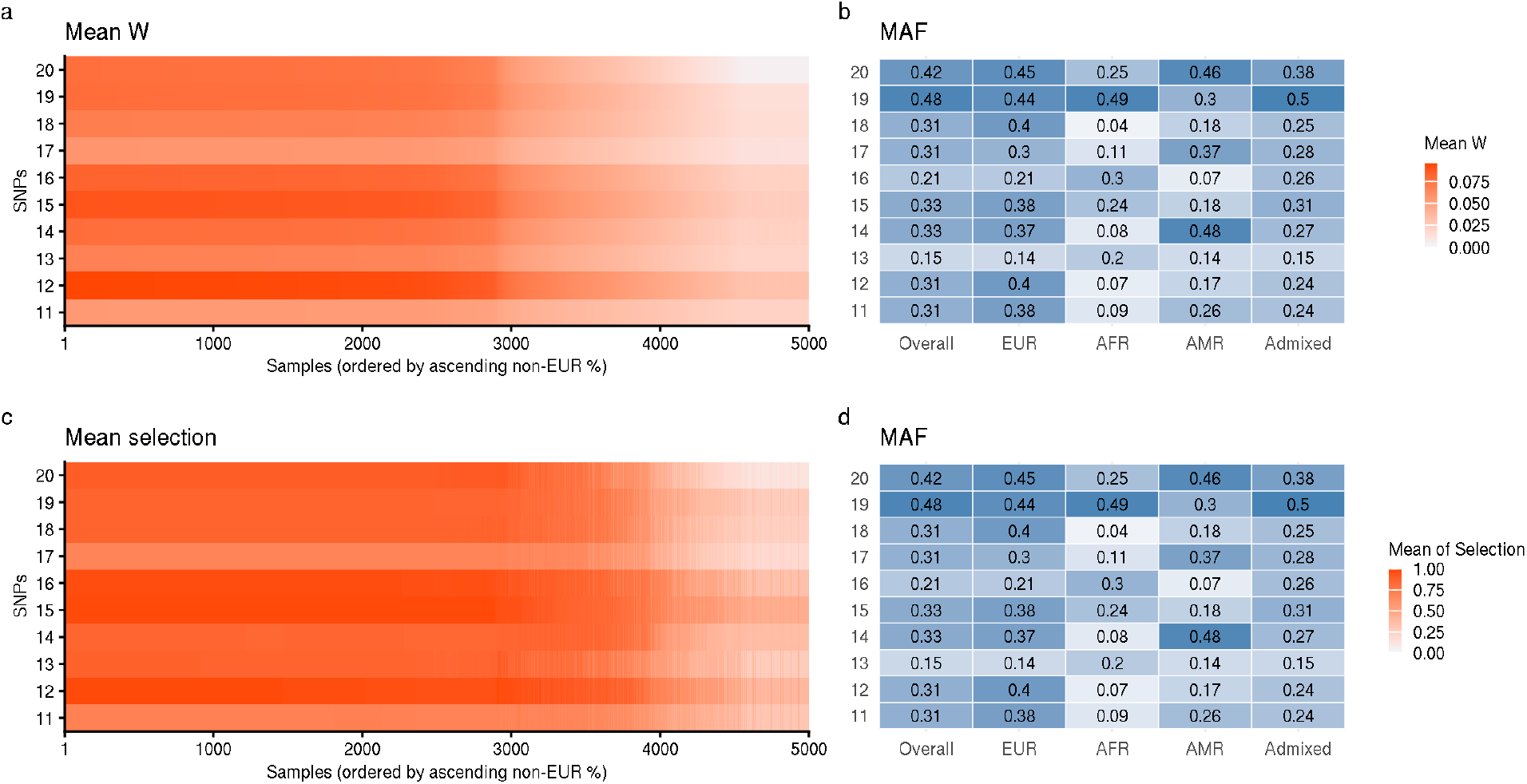
Minor allele frequencies (MAF) of EUR-interacting causal variants across stratified groups and the full samples. In panels a-d, the Y-axis corresponds to the 10 EUR-interacting causal variants used in the simulation settings described in Section 2.3. Panels **a** and **c** are the results shown in **Figure 2**, panels e and h. Panels **b** and **d** display the minor allele frequencies (MAF) of these causal SNPs across different ancestry groups and in the overall sample. Groups labeled in X-axis of b and d are defined by the estimated ancestry proportion greater than 75%; individuals not meeting this threshold are categorized as admixed.

**Figure S3.**
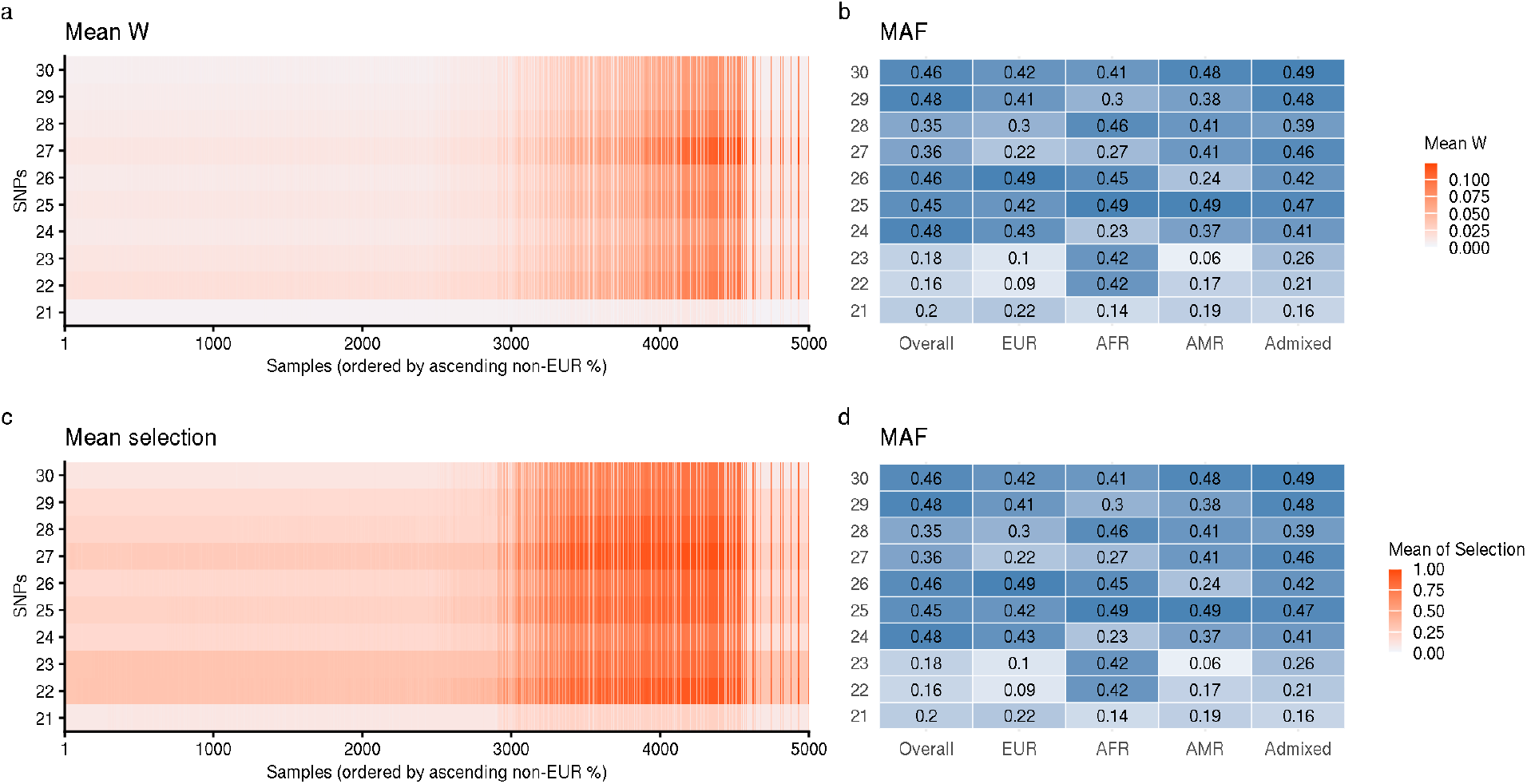
Minor allele frequencies (MAF) of AFR-interacting causal variants across stratified groups and the full samples. In panels a-d, the Y-axis corresponds to the 10 AFR-interacting causal variants used in the simulation settings described in Section 2.3. Panels **a** and **c** are the results shown in **Figure 2**, panels f and i. Panels **b** and **d** display the minor allele frequencies (MAF) of these causal SNPs across different ancestry groups and in the overall sample. Groups labeled in X-axis of b and d are defined by the estimated ancestry proportion greater than 75%; individuals not meeting this threshold are categorized as admixed.

**Figure S4.**
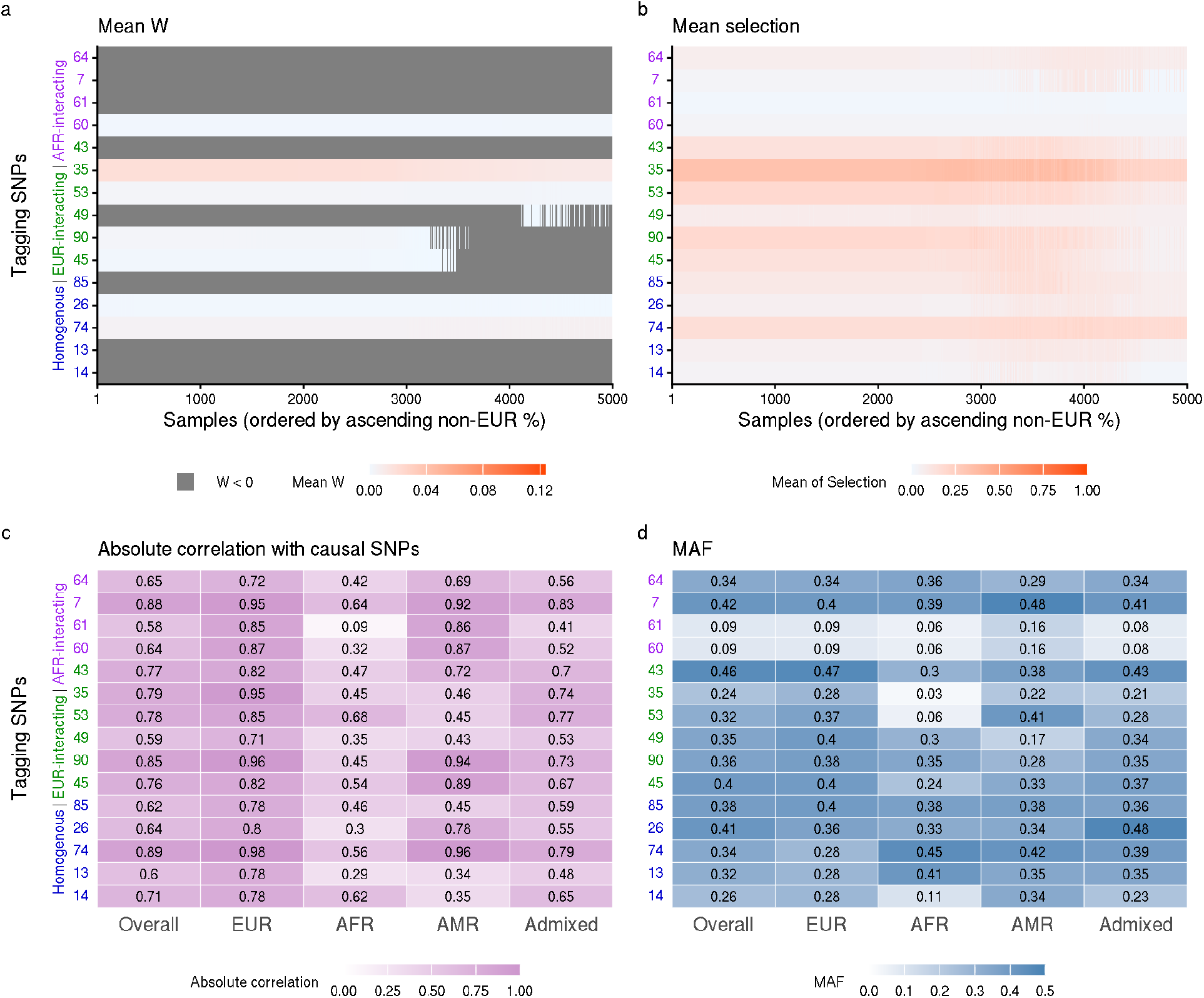
Simulation results of scenarios with both causal and tagging SNPs included in the dataset. We explored simulation results in section 3.2 to understand how PFstatistics handles linkage disequilibrium between variants. Here, tagging SNPs are defined as the top 15 non-causal variants with the highest absolute correlation to any of the 30 causal SNPs in the sample dataset, including four correlated with homogeneous SNP effects, six with EUR-interacting SNP effects, and four with AFR-interacting SNP effects. In panels a-d, the Y-axis numbers, labels and colors denote SNP indices and their corresponding groups. **a**. The mean W-statistics of tagging SNPs across 100 simulation runs were low. **b**. Tagging SNPs were rarely selected by PFstatistics when causal variants were present. **c**. The correlations between tagging and causal SNPs are shown overall and within stratified groups. **d**. Minor allele frequencies (MAF) of tagging SNPs are displayed across stratified groups and the full sample. Groups labeled in X-axis of c and d are defined by the estimated ancestry proportion greater than 75%; individuals not meeting this threshold are categorized as admixed.

**Figure S5.**
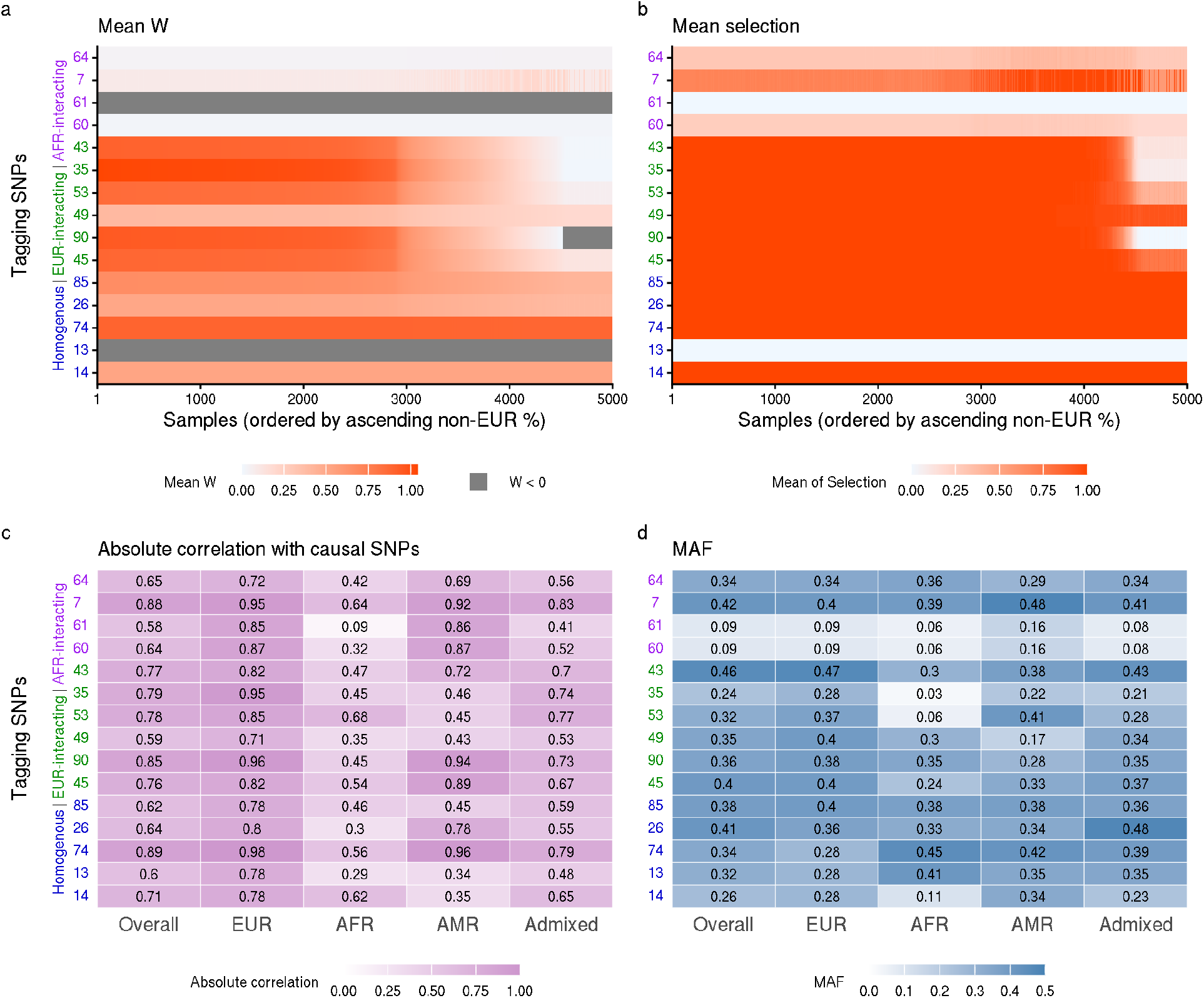
Simulation results of scenarios with causal SNPs excluded and tagging SNPs included in the dataset. We run PFstatistics on a dataset where the 30 causal SNPs used in section 3.2 were excluded from dataset, but their tagging SNPs remain. Again, tagging SNPs are defined as the top 15 non-causal variants with the highest absolute correlation to any of the 30 causal SNPs in the samples, including four correlated with homogeneous SNP effects, six with EUR-interacting SNP effects, and four with AFR-interacting SNP effects. In panels a-d, the Y-axis numbers, numbers, labels and colors denote SNP indices and their corresponding groups. **a**. The mean W-statistics of tagging SNPs resemble the trends of their corresponding causal SNPs. For example, tagging SNPs correlated with EUR-interacting variants show a gradual decrease as the proportion of non-EUR ancestry increases. **b**. Tagging SNPs became more frequent to be selected by PFstatistics when causal variants were absent, and the selection decisions exhibit similar overall patterns to those observed for causal SNPs. **c**. The correlations between tagging and causal SNPs are shown overall and within stratified groups. **d**. Minor allele frequencies (MAF) of tagging SNPs are displayed across stratified groups and the full sample. Groups labeled in X-axis of c and d are defined by the estimated ancestry proportion greater than 75%; individuals not meeting this threshold are categorized as admixed.

**Figure S6.**
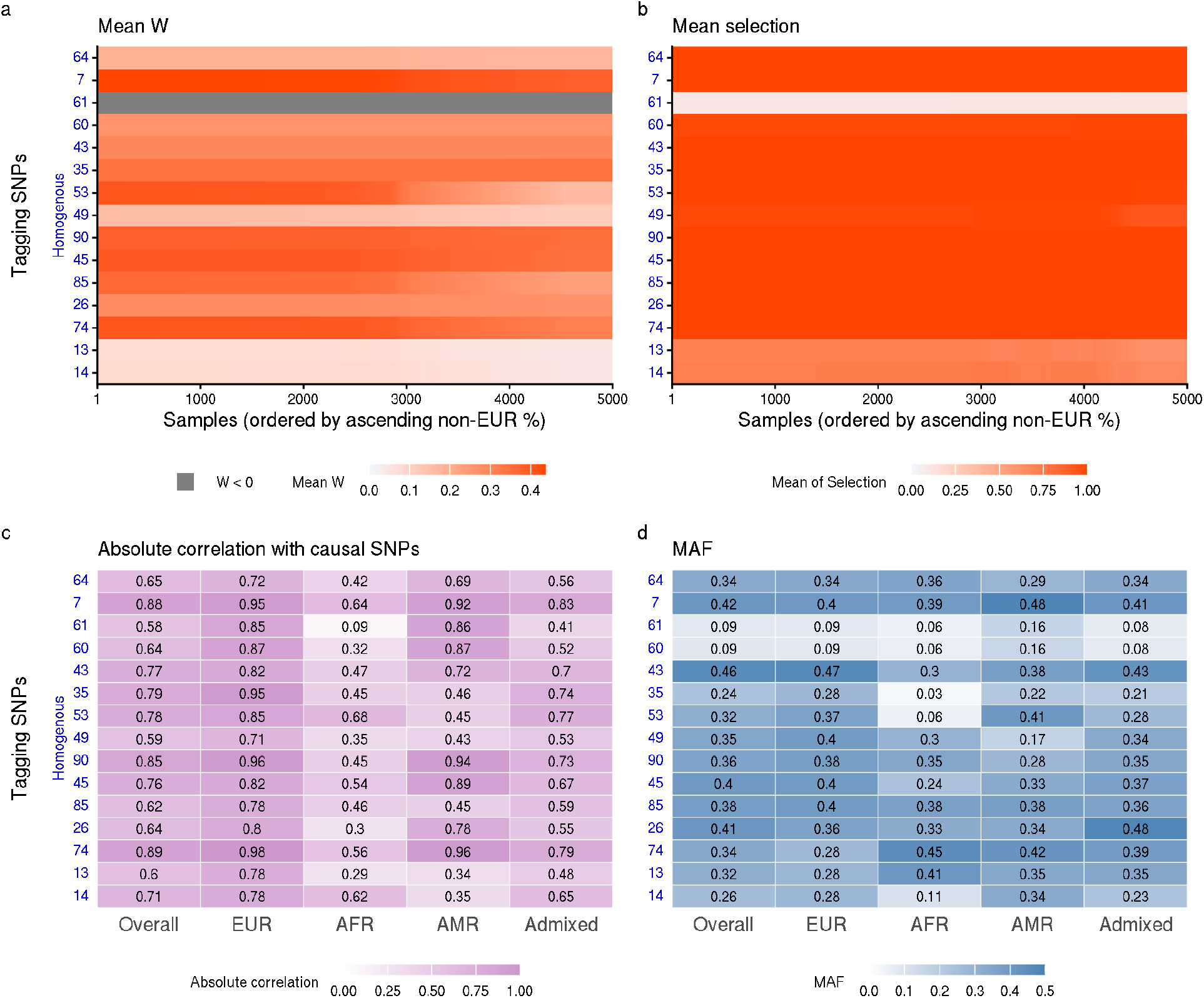
Simulation results of scenarios with 30 homogeneous causal SNPs excluded and tagging SNPs included in the dataset. We simulated phenotypes using the same 30 causal SNPs defined as in previous settings but restricted their effects to be homogeneous. We applied PFstatistics to a dataset where the 30 SNPs are excluded. Again, tagging SNPs are defined as the top 15 non-causal variants with the highest absolute correlation to any of the 30 homogenous causal SNPs in the sample dataset. In panels a-d, the Y-axis numbers, labels and colors denote SNP indices and their correspondence to homogeneous causal SNPs. **a**. The mean W-statistics of tagging SNPs resemble the trends of their corresponding causal SNPs. **b**. Tagging SNPs tend to be selected by PFstatistics when causal variants were absent, and the selection decisions exhibit similar overall patterns to those observed for causal SNPs. **c**. The correlations between tagging and causal SNPs are shown overall and within stratified groups. **d**. Minor allele frequencies (MAF) of tagging SNPs are displayed across stratified groups and the full sample. Groups labeled in X-axis of c and d are defined by the estimated ancestry proportion greater than 75%; individuals not meeting this threshold are categorized as admixed.

**Figure S7.**
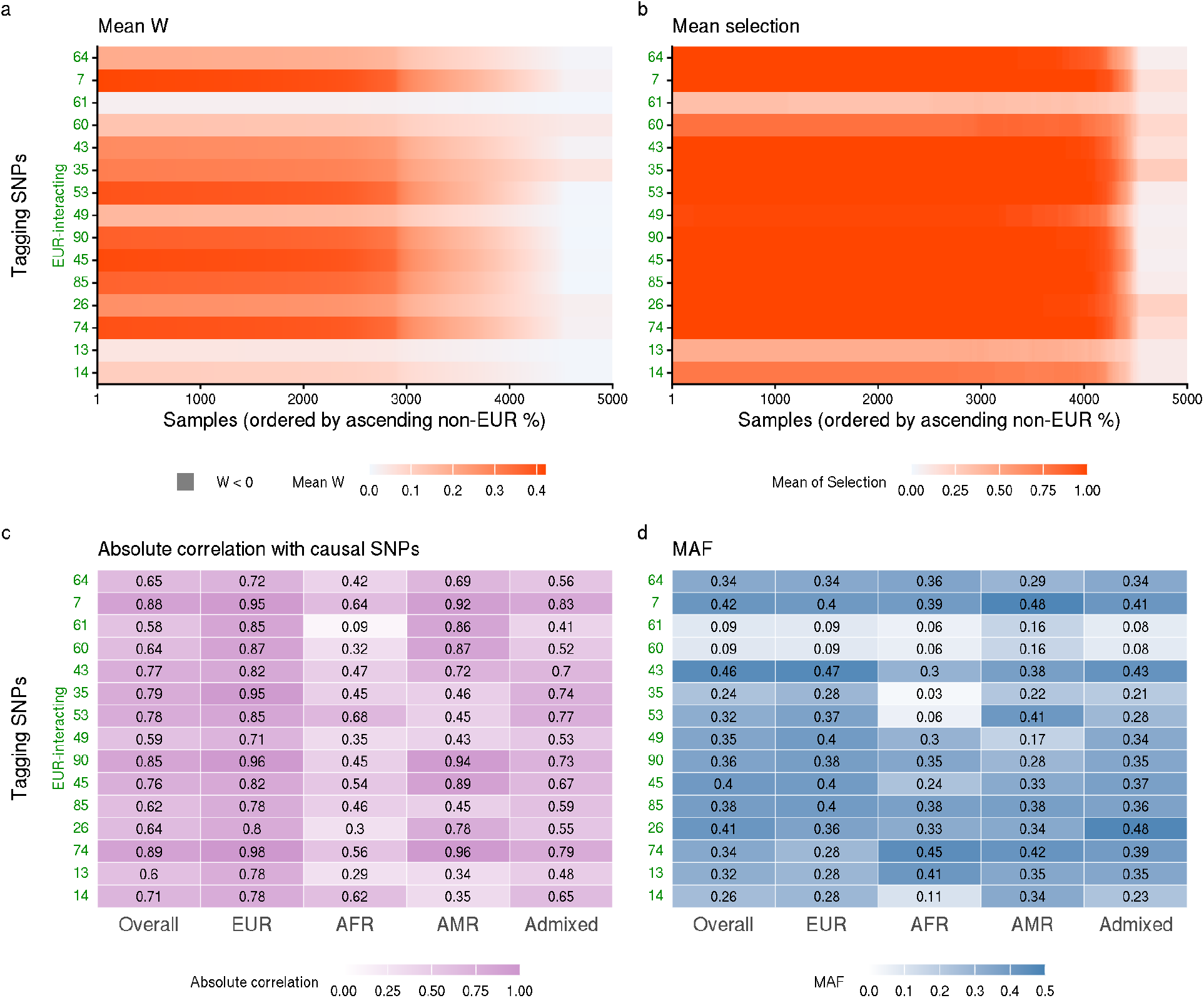
Simulation results of scenarios with 30 EUR-interacting causal SNPs excluded and tagging SNPs included in the dataset. We simulated phenotypes using the same 30 causal SNPs defined as in previous settings but restricted their effects to be interacting with EUR ancestry proportion. We applied PFstatistics to a dataset where the 30 SNPs are excluded. Again, tagging SNPs are defined as the top 15 non-causal variants with the highest absolute correlation to any of the 30 EUR-interacting causal SNPs in the sample dataset. In panels a-d, the Y-axis numbers, labels and colors denote SNP indices and their correspondence to homogeneous causal SNPs. **a**. The mean W-statistics of tagging SNPs resemble the trends of their corresponding causal SNPs. **b**. Tagging SNPs tend to be selected by PFstatistics when causal variants were absent, and the selection decisions exhibit similar overall patterns to those observed for causal SNPs. **c**. The correlations between tagging and causal SNPs are shown overall and within stratified groups. **d**. Minor allele frequencies (MAF) of tagging SNPs are displayed across stratified groups and the full sample. Groups labeled in X-axis of c and d are defined by the estimated ancestry proportion greater than 75%; individuals not meeting this threshold are categorized as admixed.

**Figure S8.**
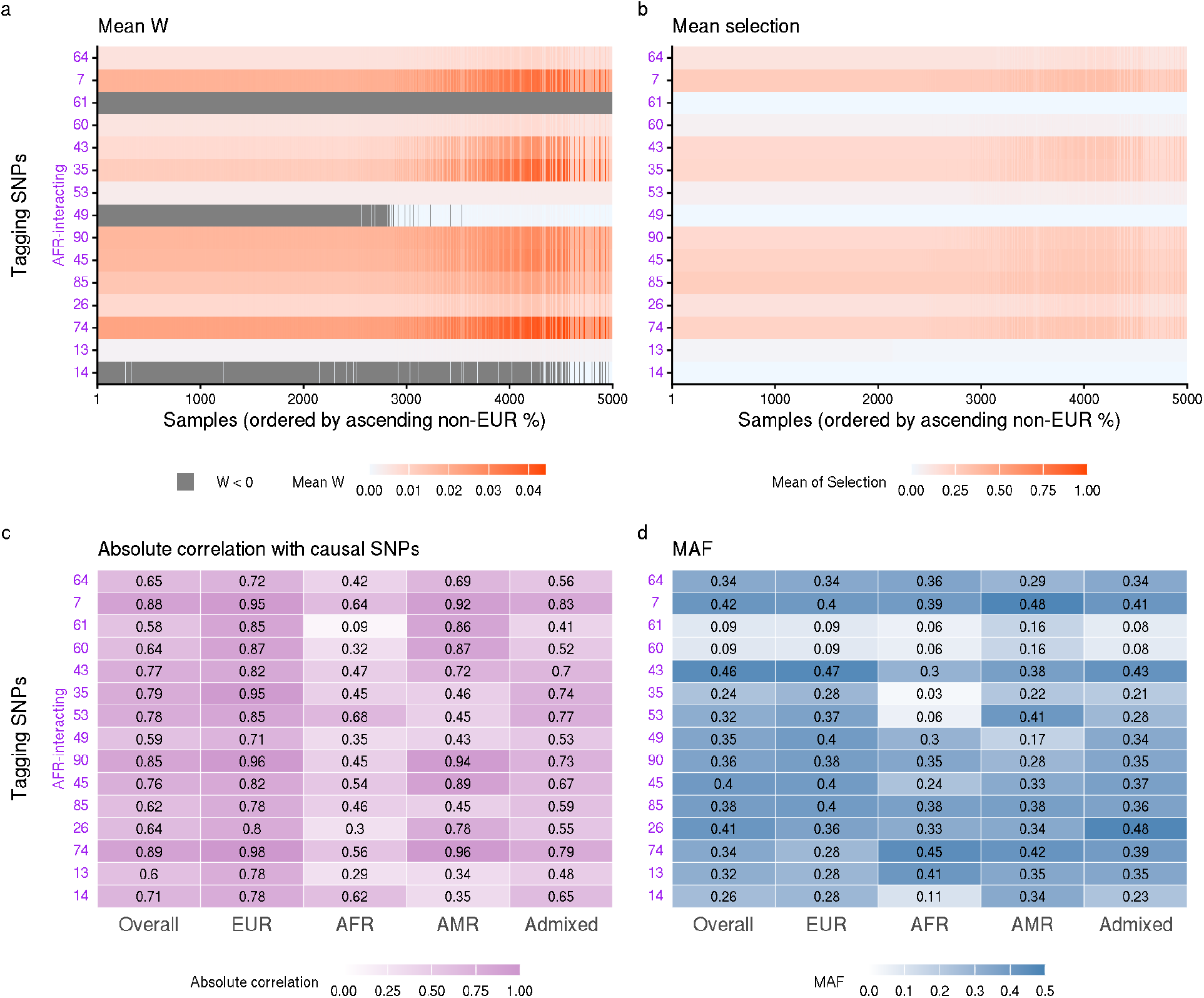
Simulation results of scenarios with 30 AFR-interacting causal SNPs excluded and tagging SNPs included in the dataset. We simulated phenotypes using the same 30 causal SNPs defined as in previous settings but restricted their effects to be interacting with the estimated AFR ancestry proportion. We applied PFstatistics to a dataset where the 30 SNPs are excluded. Again, tagging SNPs are defined as the top 15 non-causal variants with the highest absolute correlation to any of the 30 AFR-interacting causal SNPs in the sample dataset. In panels a-d, the Y-axis numbers, labels and colors denote SNP indices and their correspondence to homogeneous causal SNPs. **a**. The mean W-statistics of tagging SNPs resemble the trends of their corresponding causal SNPs. **b**. Tagging SNPs tend to be selected by PFstatistics when causal variants were absent, and the selection decisions exhibit similar overall patterns to those observed for causal SNPs. **c**. The correlations between tagging and causal SNPs are shown overall and within stratified groups. **d**. Minor allele frequencies (MAF) of tagging SNPs are displayed across stratified groups and the full sample. Groups labeled in X-axis of c and d are defined by the estimated ancestry proportion greater than 75%; individuals not meeting this threshold are categorized as admixed.

**Figure S9.**
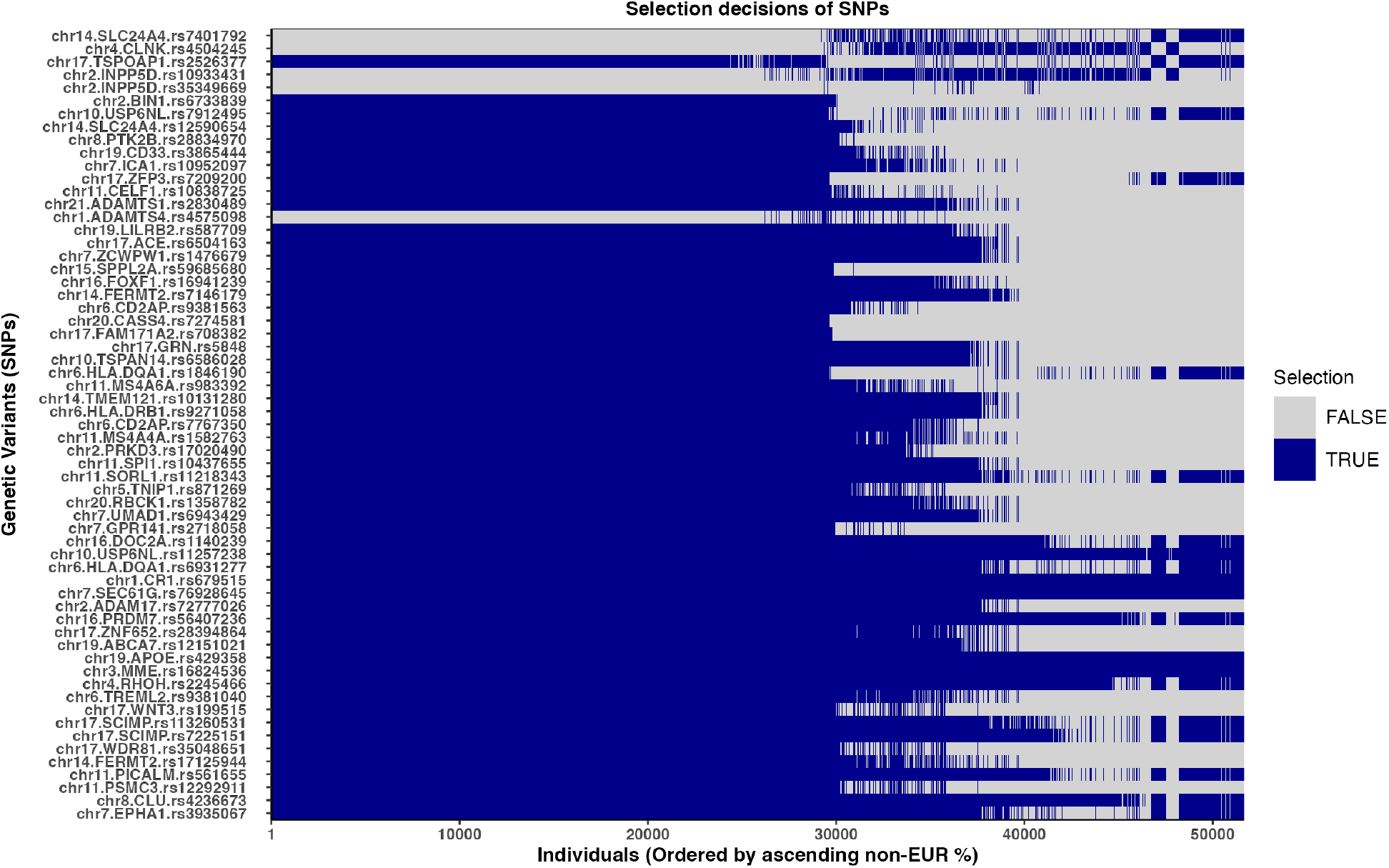
Individual-level variant selection matrix generated by PFstatistics on ADSP data. The matrix highlights heterogeneous selection patterns across individuals for SNPs selected in at least one individual in the cohort, same as SNPs present in Figure 3g. The ordering of these SNPs matches that of Figure 3g.

**Figure S10.**
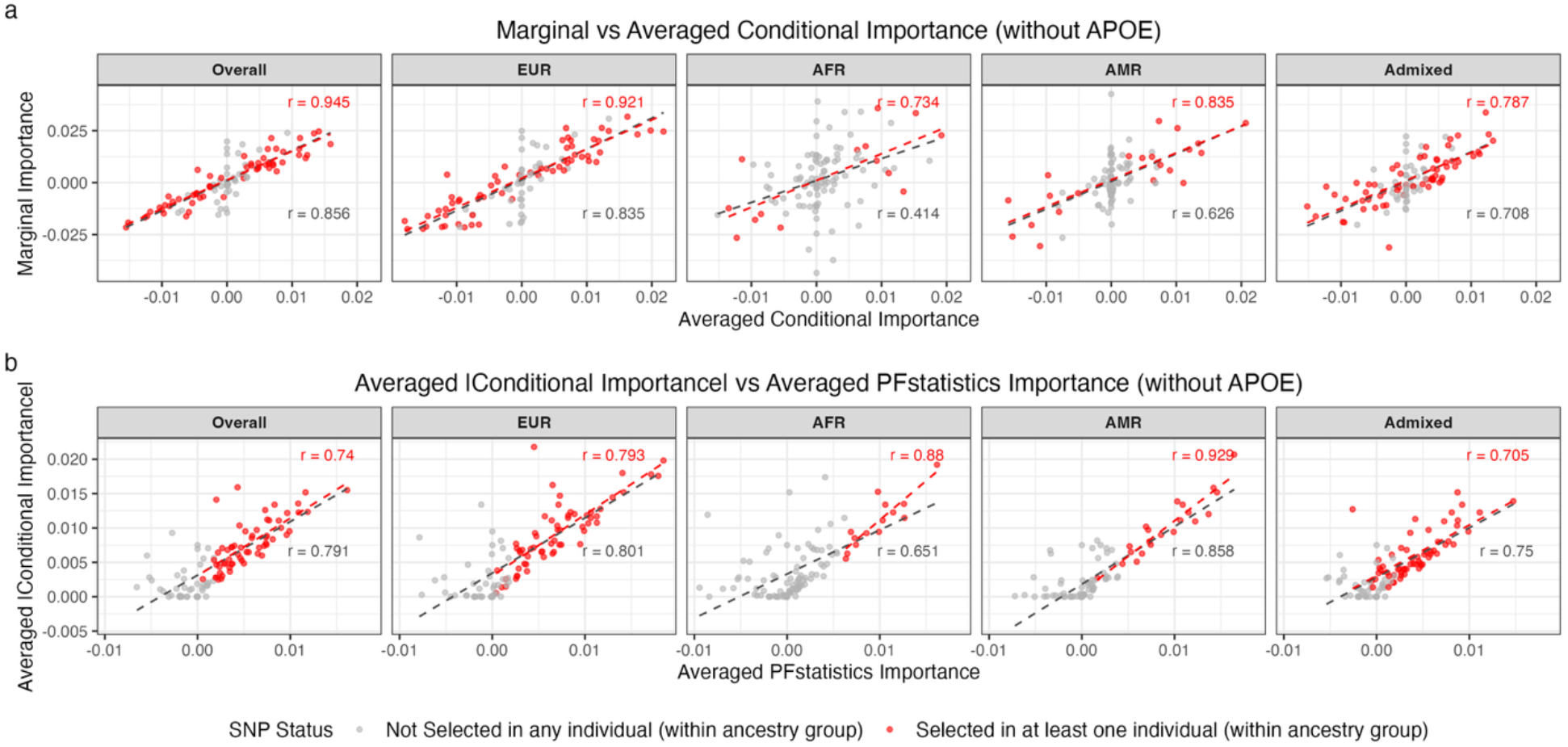
Comparison between marginal, conditional, and PFstatistics importance scores across ancestry groups (without APOE) **a**. Scatterplots comparing marginal importance scores from ancestry-stratified Gaussian regression models with conditional importance scores derived from the Lasso interaction model, shown separately for the Overall sample and for each ancestry group (EUR, AFR, AMR, and Admixed). **b**. Scatterplots comparing in stratified groups the averaged absolute conditional importance scores with ancestry-averaged knockoff importance scores (W-statistics) produced by PFstatistics for the same SNP set. In both panels, each point represents a SNP. Red points indicate SNPs selected in at least one individual by PFstatistics within the corresponding ancestry group, whereas gray points represent SNPs not selected by PFstatistics in any individuals in that group. Red dashed lines represent the regression fitted to the selected SNPs (red points) within each ancestry group, while the gray dashed line represents the regression fitted to all SNPs. Pearson correlation coefficients (r) are reported for selected SNPs (red texts) and for all SNPs (grey texts), quantifying agreement between importance measures within each ancestry stratum.

## Supplemental Notes: An illustrative application to polygenic risk score (PRS) construction by leveraging heterogeneous genetic effects

Substantial efforts have been made to develop PRS, but most existing models are primarily trained on European-ancestry populations, limiting their transferability and predictive accuracy in non-European populations. The heterogeneous genetic effects identified by PFstatistics naturally allow us to incorporate this information into PRS construction. Although the focus of this paper is on variable selection instead of developing PRS, we present a proof-of-concept example to showcase how one can improve existing PRS by leveraging heterogeneous genetic effects.

Our proposed PFstatistics framework can be naturally extended to construct a PRS to incorporate heterogeneous genetic effects by modeling both main genetic effects and interactions with heterogeneity features. We define our PFstatistics PRS as:

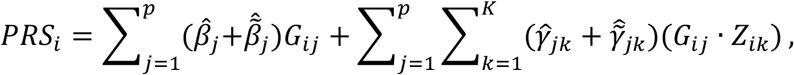

where 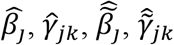 are estimated effect sizes corresponding to genotype and interacted effects of original and knockoff SNPs estimated from our training model in **Eq. 3**. We substitute knockoff variables with the original variables, as knockoff generation introduces inherent randomness. Since the effect sizes of knockoff variables are largely independent of the outcome, their estimated coefficients typically shrink toward zero, contributing minimally to the PRS. The continuous heterogeneous genetic effects captured by PFstatistics suggest a potential role for such information in PRS construction. We evaluated our PFstatistics-based PRS performance under simulation settings and in real data analysis, compared with four common PRS constructions:

a. *Full-Sample Marginal Test PRS* estimates SNP effects by fitting a separate linear regression of the phenotype on each SNP using all samples, adjusting for covariates. For each SNP, we obtain an effect size estimate which is used as the SNP weight. An individual’s PRS is computed as the weighted sum of their genotypes across all SNPs.
b. *Full-Sample Lasso PRS* is trained on the full, ancestry-inclusive sample using a penalized regression model that includes both genotypes and covariates as predictors. We fit a lasso model with 10-fold cross-validation to select the tuning parameter, while setting penalty factors to zero for covariates and one for SNPs to ensure that only SNP effects are penalized. The model is implemented using the cv.glmnet function from the R package glmnet. PRS values for test individuals are computed as the linear combination of each individual’s SNP genotypes with their estimated lasso coefficients.
c. *EUR-trained Lasso PRS* follows the same lasso regression framework, but the model is trained exclusively on individuals of European ancestry in the training data. The fitted coefficients are then applied to compute PRS for the target validation populations, which may include EUR, AFR, AMR, or admixed individuals. This setup mimics the common scenario in which PRS models are trained on large-scale European datasets and then transferred to non-European populations without re-estimation.
d. Analogously, *AFR-trained Lasso PRS* is trained solely on individuals of African ancestry in the training data, using the same penalized regression framework. The estimated coefficients are then used to compute PRS in the multi-ancestry validation set.

We first evaluated PRS performance in a simulation study designed to mimic ancestry-dependent genetic effects using real genotype data from the ADSP (**Figure S11a**). A random subset of 10,000 individuals was split into training (n = 5,000) and test (n = 5,000) sets, and variants with minor allele frequency (MAF) below 5% in the subset were excluded. True genetic effects were generated to vary continuously across ancestry by assigning subsets of SNPs effects that depended on individual-level European (EUR) and African (AFR) ancestry proportions, in addition to homogeneous baseline effects. Outcomes were simulated as the sum of these scaled genetic effects and independent Gaussian noise, with noise resampled in each replicate. We repeated the entire procedure 100 times and evaluated each PRS method in the full test set and within ancestry-stratified subgroups (EUR, AFR, AMR, and admixed), using AUC as the performance metric. Across all groups, PRS constructed from PFstatistics showed the strongest and most stable overall performance, consistently outperforming PRS based on full-sample marginal association tests and full-sample lasso fits. PFstatistics PRS was comparable to ancestry-matched lasso PRS when evaluated within the corresponding ancestry group, such as PFstatistics PRS versus EUR-trained lasso PRS in EUR individuals and PFstatistics PRS versus AFR-trained lasso PRS in AFR individuals. However, ancestry-specific lasso PRS generalized poorly outside the ancestry group on which they were trained. In contrast, PFstatistics PRS maintained comparatively robust performance across all groups, consistent with its ability to model ancestry as a continuous source of effect heterogeneity rather than relying on discrete ancestry-specific training sets.

We next assessed these PRS methods in the ADSP data using nested cross-validation (**Figure S11b–f**). The full dataset was partitioned into five outer folds, each used once as a held-out test fold, while the remaining samples formed the corresponding training set. Within each outer training split, five-fold inner cross-validation was used to tune model hyperparameters, after which the final model was refit on the complete outer-fold training data and applied to the held-out outer-fold test set. Heterogeneity features, including ancestry-related covariates and principal components, were included as unpenalized predictors in all relevant models. For primary prediction analyses, performance was summarized using pooled AUC computed from predictions aggregated across all five outer test folds, both overall and within ancestry-stratified groups (**Figure S11b**). PFstatistics PRS did not provide a substantial improvement in overall case-control discrimination relative to simpler PRS approaches based on full-sample marginal tests or lasso, and most methods showed broadly similar AUC across groups.

To further characterize clinical utility beyond overall discrimination, we evaluated the relationship between PRS and age at onset (AAO) among individuals with MCI or AD (**Figure S11c**). Across most ancestry groups, higher PRS tended to be associated with earlier onset, as reflected by negative values of the Spearman correlation between PRS and AAO. This pattern was strongest in AFR and more modest in the other groups, whereas associations in AMR were weak across methods. We also examined the pooled distribution of PRS scores in cases and controls (**Figure S11d**), which showed a rightward shift among cases relative to controls but substantial overlap between the two distributions, consistent with the modest overall discrimination observed in panel b.

Because PRS may be most informative at the extremes of the risk distribution, we next restricted evaluation to individuals in the top and bottom 10% of the pooled PRS distribution (**Figure S11e, f**). In this extreme-risk setting, discrimination improved for all methods relative to the full-sample analysis, with pooled AUC values notably higher across the overall sample and most ancestry groups (**Figure S11e**). Similarly, the odds ratio comparing disease risk in the top 10% versus bottom 10% of the PRS distribution was substantially elevated across methods, particularly in EUR and, to a lesser extent, AFR (**Figure S11f**). PFstatistics PRS was among the best-performing methods in these extreme-strata analyses. Taken together, these results suggest that, in ADSP, the main advantage of PFstatistics is not a dramatic improvement in aggregate prediction accuracy, but rather its ability to provide an ancestry-aware PRS framework that remains competitive across diverse and admixed populations while naturally accommodating continuously varying genetic effects.

**Figure S11.**
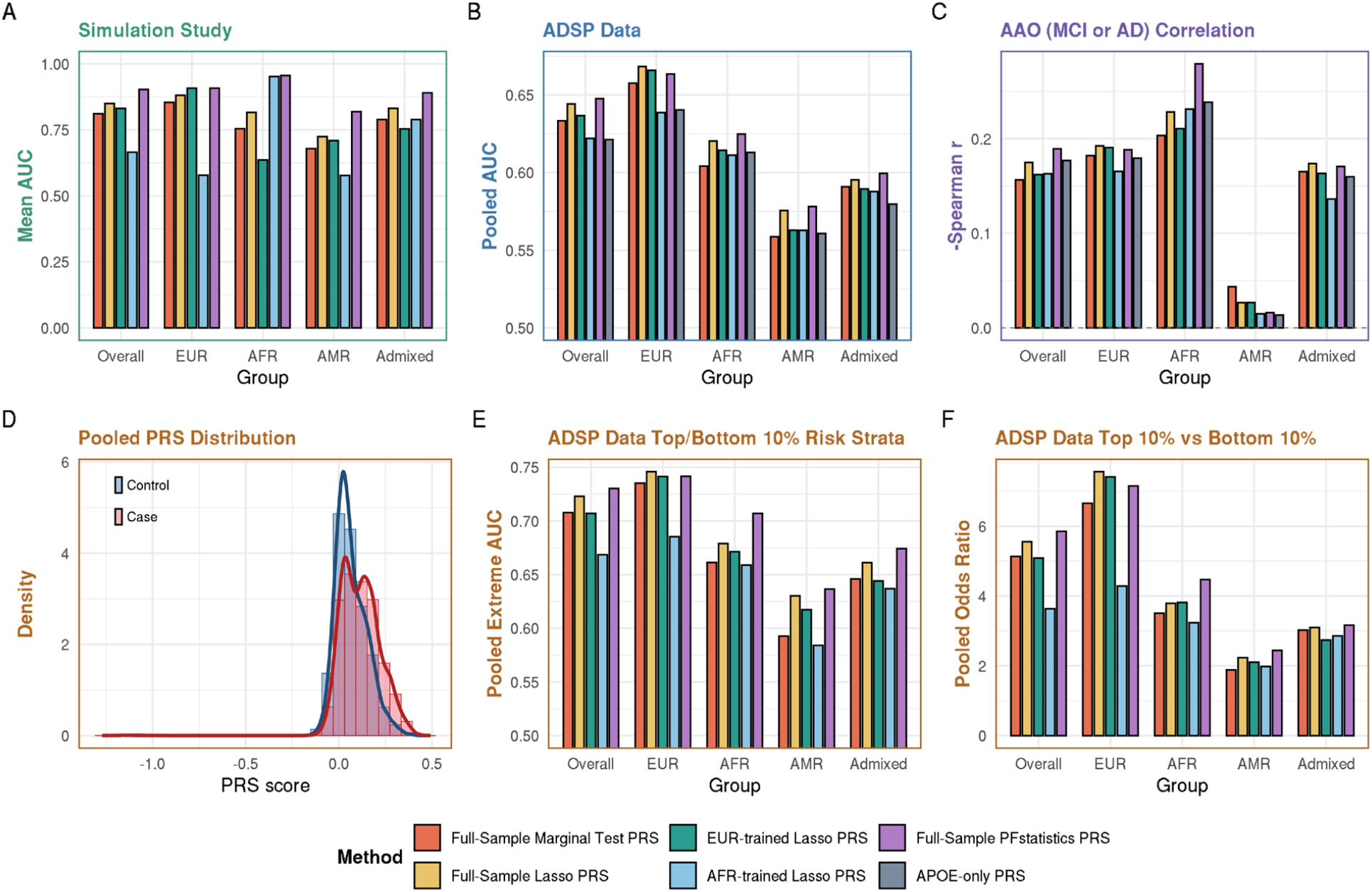
Performance of PRS methods across simulation and ADSP data. **a**. Simulation results. Bar plots show the mean AUC in left-out validation samples (n=5000). Results are stratified by overall population and by EUR, AFR, AMR, and admixed groups. **b**. ADSP data results. Bar plots show the pooled AUC based on predictions from all five outer test folds in a five-fold outer, five-fold inner nested cross-validation framework, stratified by the overall population and by EUR, AFR, AMR, and admixed groups. **c**. Association with age at onset (AAO) among individuals with MCI or AD. Bar plots show the negative Spearman correlation between PRS and AAO, stratified by ancestry group. Larger values indicate stronger association with earlier age at onset. **d**. Distribution of pooled PRS in ADSP. Histogram and density curves show the distribution of pooled PRS scores in controls and cases. **e**. Discriminative performance in the extreme-risk strata of ADSP. Bar plots show the pooled AUC when evaluation is restricted to individuals in the top 10% and bottom 10% of the PRS distribution in each ancestry group. **f**. Enrichment of disease risk in the extreme-risk strata of ADSP. Bar plots show the pooled odds ratio comparing individuals in the top 10% versus the bottom 10% of the PRS distribution, stratified by ancestry group. Colors indicate PRS construction methods: full-sample marginal test PRS, full-sample lasso PRS, EUR-trained lasso PRS, AFR-trained lasso PRS, full-sample PFstatistics PRS, and APOE-only PRS.

