## Supplemental Information for "Personalized Feature Statistics: Individual-Level Variant Inference under Genetic Ancestry Continuum"

### Supplemental Information for "Profiling Heterogeneous Genetic Effect Across Genetic Ancestry Continuum"

#### Supplemental Figures

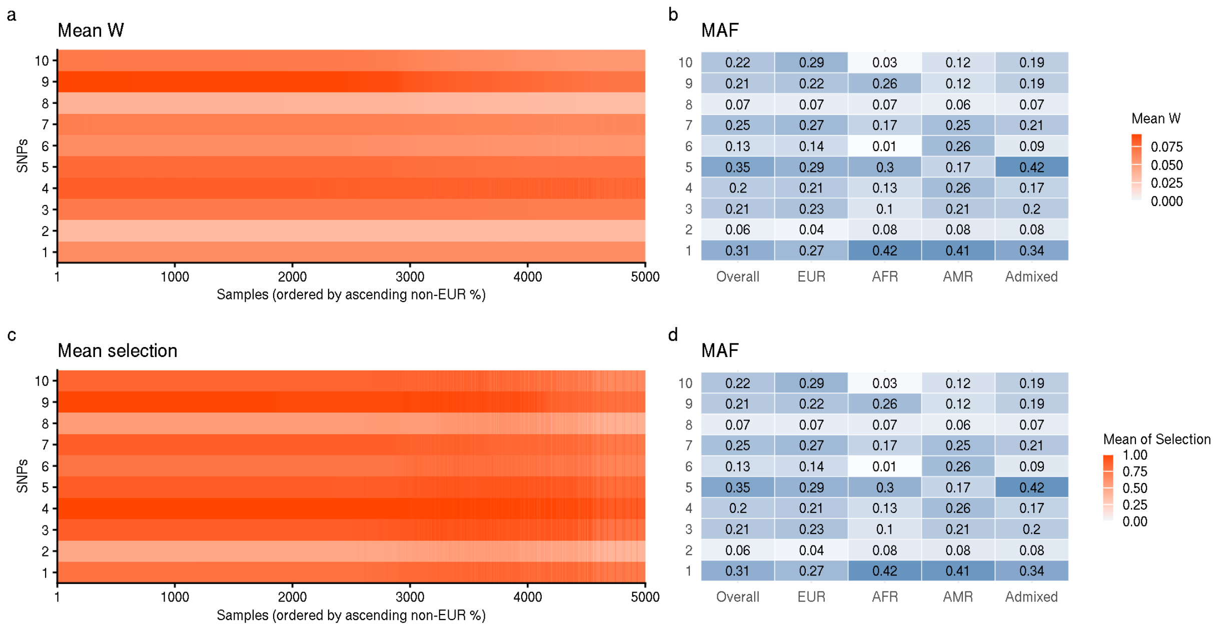


**Figure S1. Minor allele frequencies (MAF) of homogenous causal variants across stratified groups and the full samples**

In panels a-d, the Y-axis corresponds to the 10 homogeneous causal variants used in the simulation settings described in Section 2.3. Panels **a** and **c** are the results shown in **Figure 2**, panels d and g. Panels **b** and **d** display the minor allele frequencies (MAF) of these causal SNPs across different ancestry groups and in the overall sample. Groups labeled in X-axis of b and d are defined by the estimated ancestry proportion greater than 75%; individuals not meeting this threshold are categorized as admixed.

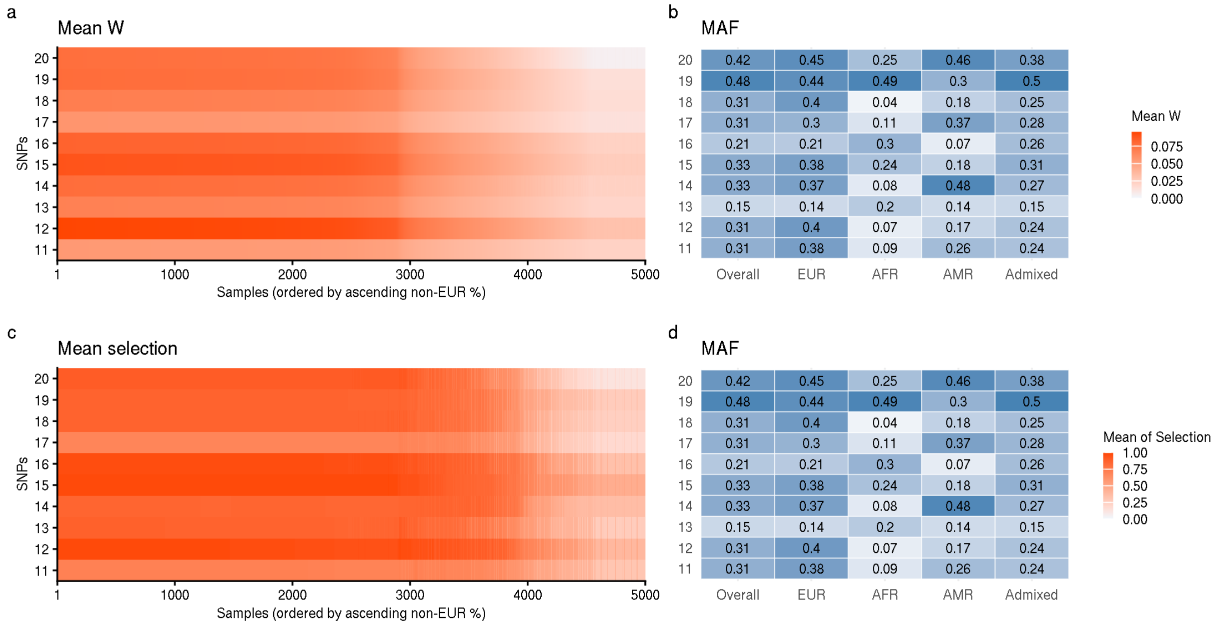


**Figure S2. Minor allele frequencies (MAF) of EUR-interacting causal variants across stratified groups and the full samples**

In panels a-d, the Y-axis corresponds to the 10 EUR-interacting causal variants used in the simulation settings described in Section 2.3. Panels **a** and **c** are the results shown in **Figure 2**, panels e and h. Panels **b** and **d** display the minor allele frequencies (MAF) of these causal SNPs across different ancestry groups and in the overall sample. Groups labeled in X-axis of b and d are defined by the estimated ancestry proportion greater than 75%; individuals not meeting this threshold are categorized as admixed.

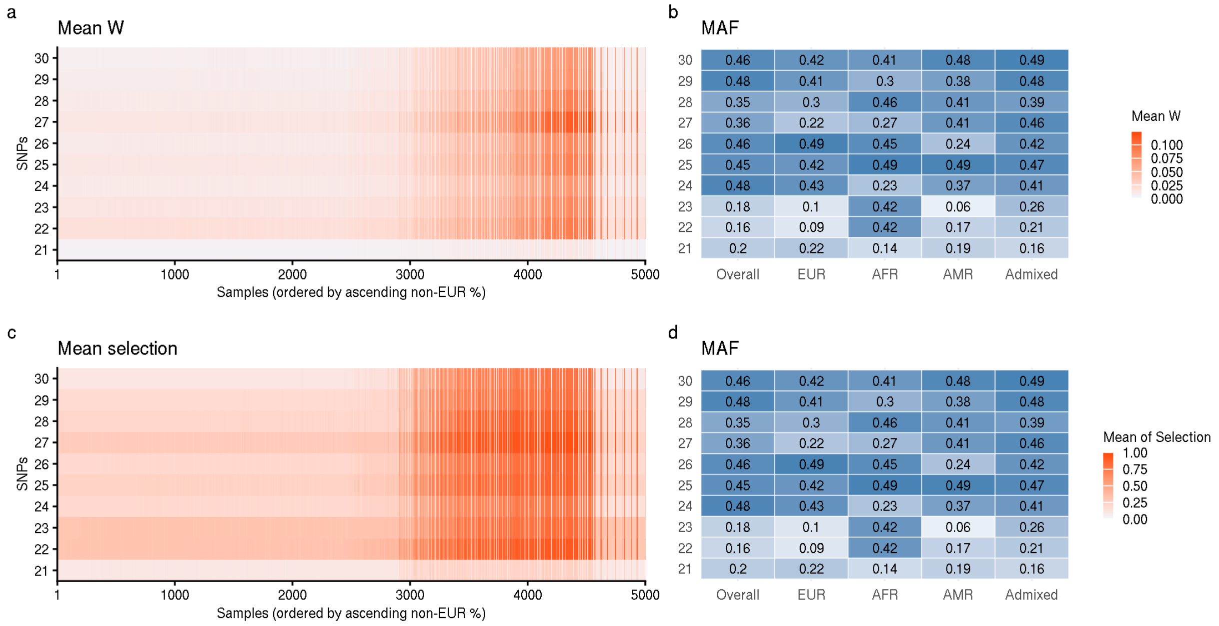


**Figure S3. Minor allele frequencies (MAF) of AFR-interacting causal variants across stratified groups and the full samples**

In panels a-d, the Y-axis corresponds to the 10 AFR-interacting causal variants used in the simulation settings described in Section 2.3. Panels **a** and **c** are the results shown in **Figure 2**, panels f and i. Panels **b** and **d** display the minor allele frequencies (MAF) of these causal SNPs across different ancestry groups and in the overall sample. Groups labeled in X-axis of b and d are defined by the estimated ancestry proportion greater than 75%; individuals not meeting this threshold are categorized as admixed.

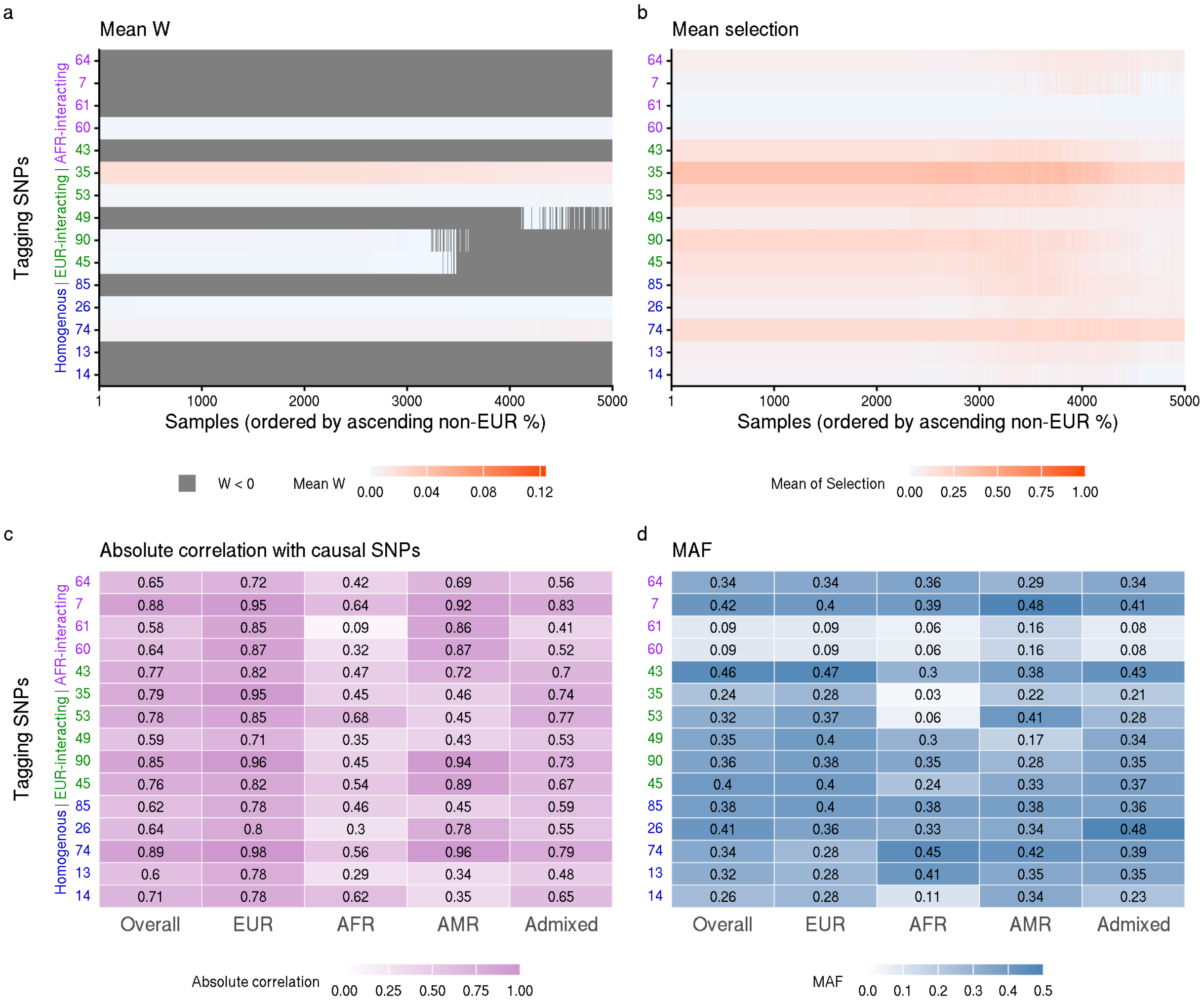


**Figure S4. Simulation results of scenarios with both causal and tagging SNPs included in the dataset**

We explored simulation results in section 3.2 to understand how PFstatistics handles linkage disequilibrium between variants. Here, tagging SNPs are defined as the top 15 non-causal variants with the highest absolute correlation to any of the 30 causal SNPs in the sample dataset, including four correlated with homogeneous SNP effects, six with EUR-interacting SNP effects, and four with AFR-interacting SNP effects. In panels a-d, the Y-axis numbers, labels and colors denote SNP indices and their corresponding groups. **a.** The mean W-statistics of tagging SNPs across 100 simulation runs were low. **b.** Tagging SNPs were rarely selected by PFstatistics when causal variants were present. **c.** The correlations between tagging and causal SNPs are shown overall and within stratified groups. **d.** Minor allele frequencies (MAF) of tagging SNPs are displayed across stratified groups and the full sample. Groups labeled in X-axis of c and d are defined by the estimated ancestry proportion greater than 75%; individuals not meeting this threshold are categorized as admixed.

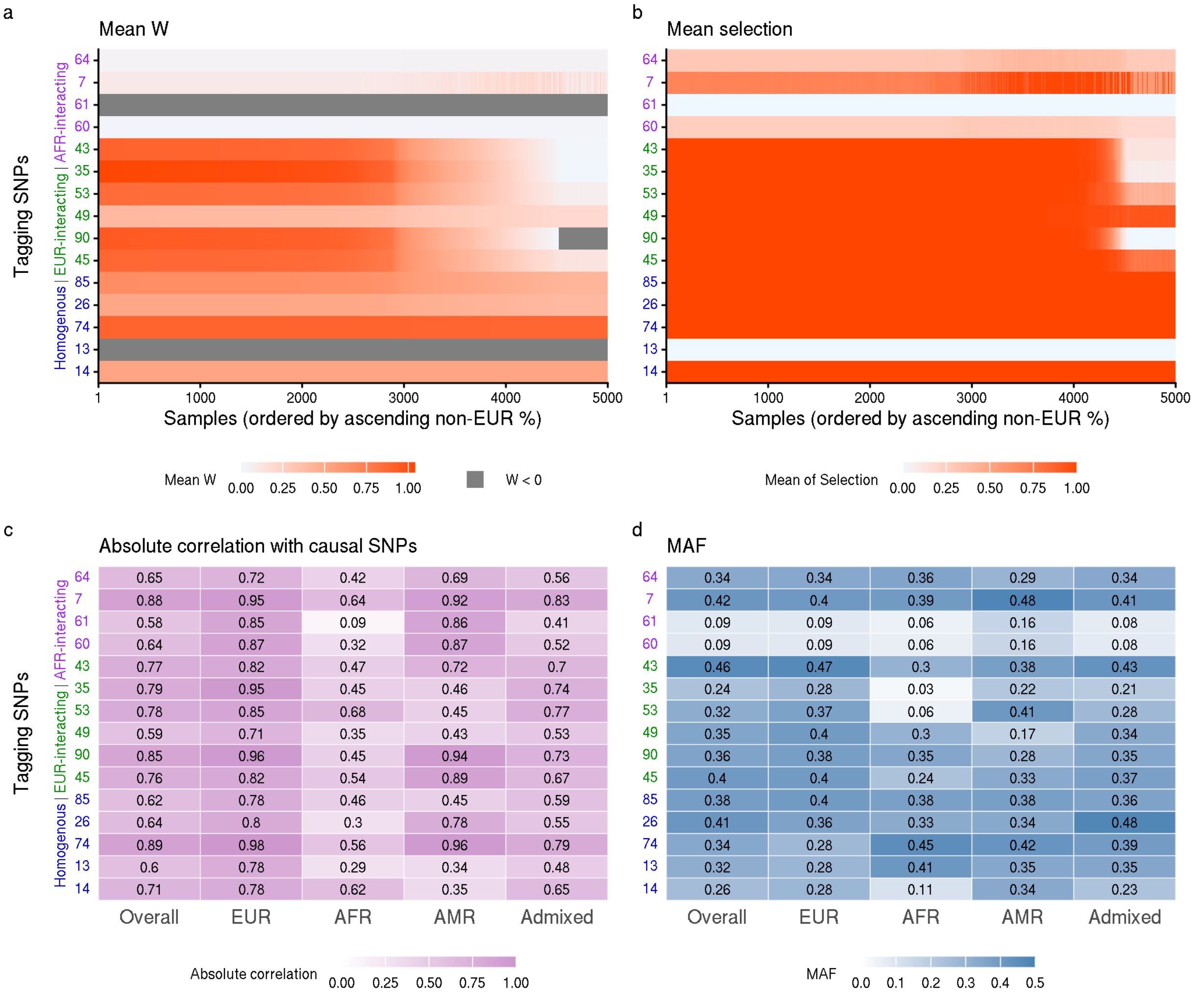


**Figure S5. Simulation results of scenarios with causal SNPs excluded and tagging SNPs included in the dataset**

We run PFstatistics on a dataset where the 30 causal SNPs used in section 3.2 were excluded from dataset, but their tagging SNPs remain. Again, tagging SNPs are defined as the top 15 non-causal variants with the highest absolute correlation to any of the 30 causal SNPs in the samples, including four correlated with homogeneous SNP effects, six with EUR-interacting SNP effects, and four with AFR-interacting SNP effects. In panels a-d, the Y-axis numbers, numbers, labels and colors denote SNP indices and their corresponding groups. **a.** The mean W-statistics of tagging SNPs resemble the trends of their corresponding causal SNPs. For example, tagging SNPs correlated with EUR-interacting variants show a gradual decrease as the proportion of non-EUR ancestry increases. **b.** Tagging SNPs became more frequent to be selected by PFstatistics when causal variants were absent, and the selection decisions exhibit similar overall patterns to those observed for causal SNPs. **c.** The correlations between tagging and causal SNPs are shown overall and within stratified groups. **d.** Minor allele frequencies (MAF) of tagging SNPs are displayed across stratified groups and the full sample. Groups labeled in X-axis of c and d are defined by the estimated ancestry proportion greater than 75%; individuals not meeting this threshold are categorized as admixed.

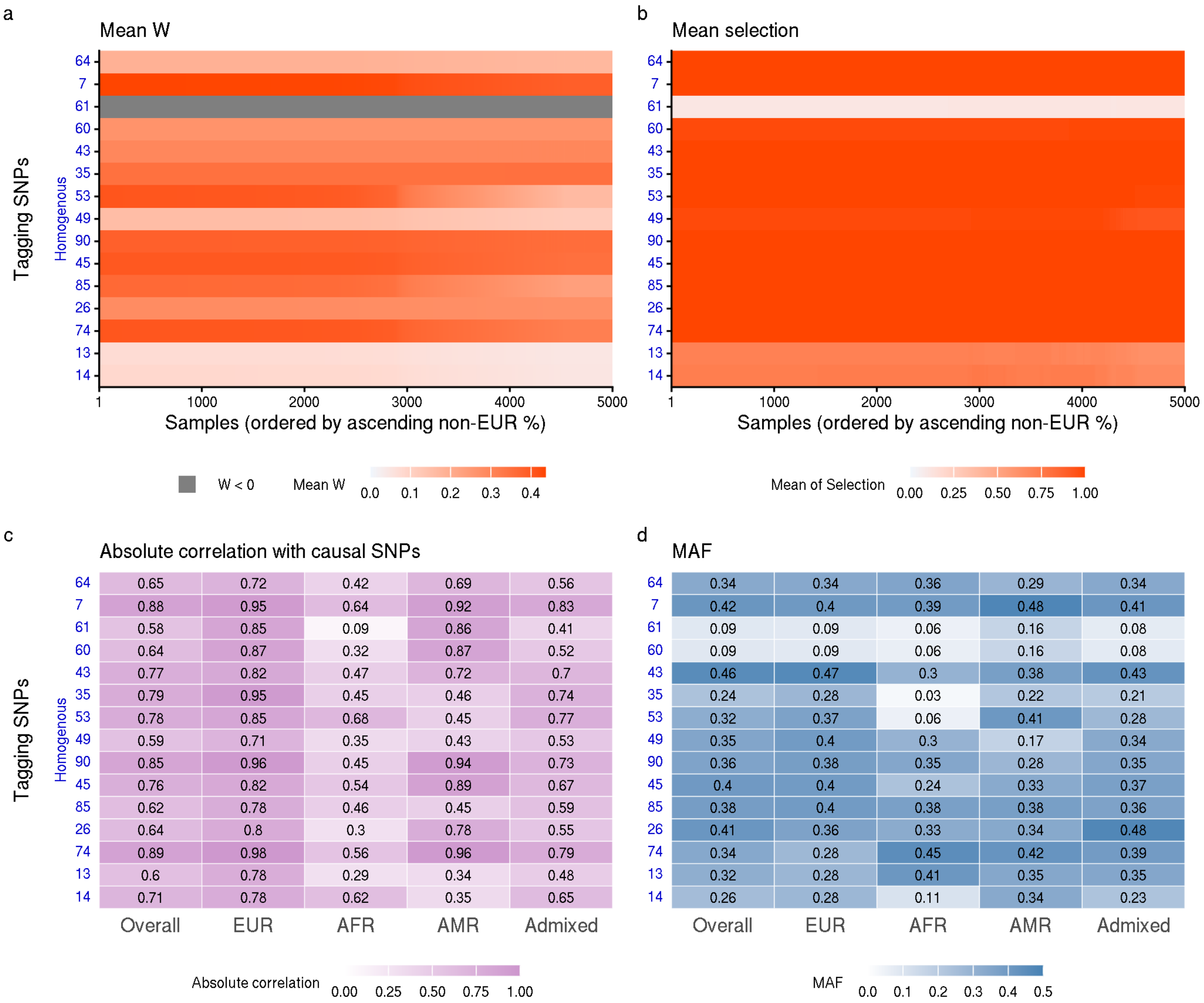


**Figure S6. Simulation results of scenarios with 30 homogeneous causal SNPs excluded and tagging SNPs included in the dataset**

We simulated phenotypes using the same 30 causal SNPs defined as in previous settings but restricted their effects to be homogeneous. We applied PFstatistics to a dataset where the 30 SNPs are excluded. Again, tagging SNPs are defined as the top 15 non-causal variants with the highest absolute correlation to any of the 30 homogenous causal SNPs in the sample dataset. In panels a-d, the Y-axis numbers, labels and colors denote SNP indices and their correspondence to homogeneous causal SNPs. **a.** The mean W-statistics of tagging SNPs resemble the trends of their corresponding causal SNPs. **b.** Tagging SNPs tend to be selected by PFstatistics when causal variants were absent, and the selection decisions exhibit similar overall patterns to those observed for causal SNPs. **c.** The correlations between tagging and causal SNPs are shown overall and within stratified groups. **d.** Minor allele frequencies (MAF) of tagging SNPs are displayed across stratified groups and the full sample. Groups labeled in X-axis of c and d are defined by the estimated ancestry proportion greater than 75%; individuals not meeting this threshold are categorized as admixed.

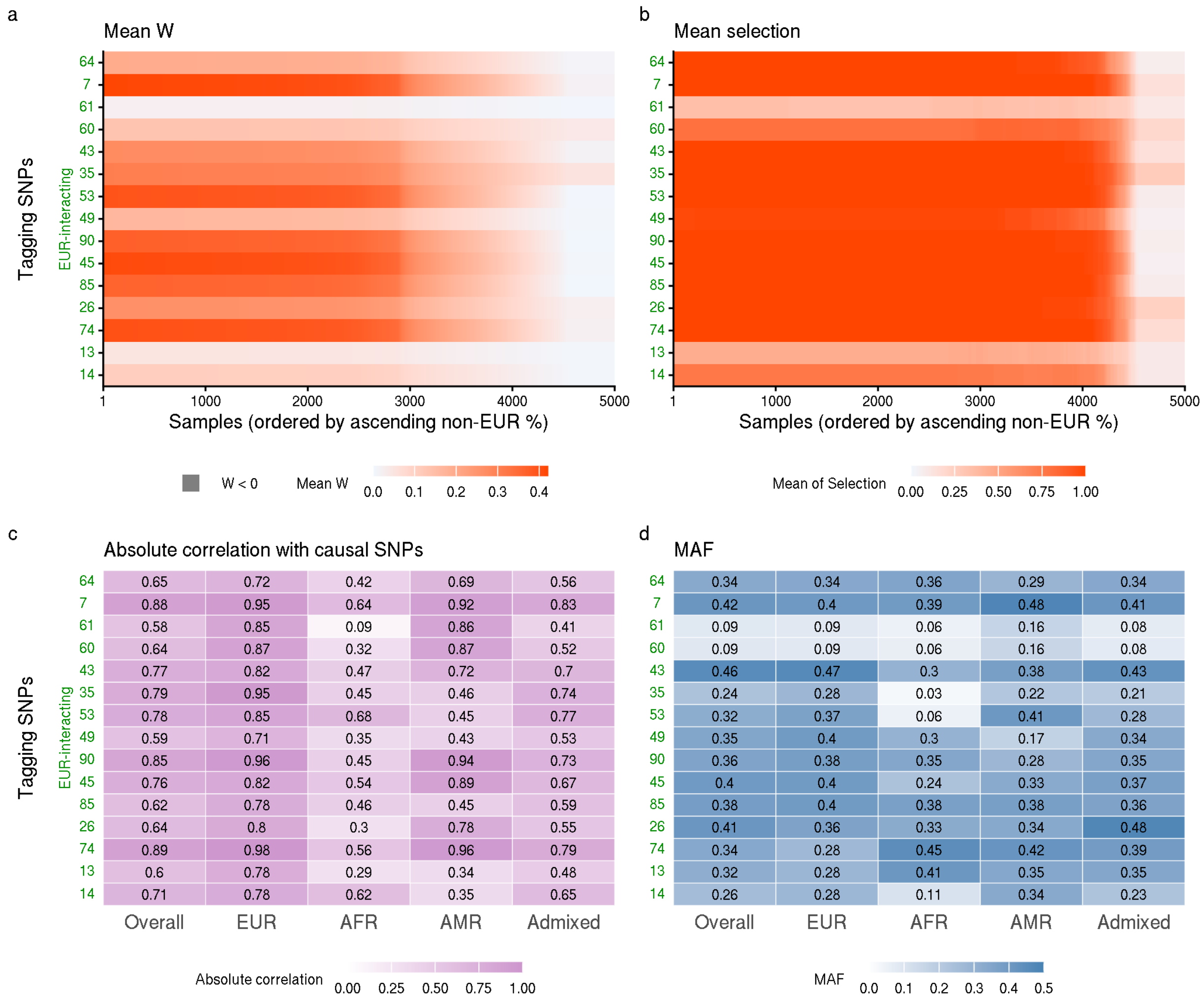


**Figure S7. Simulation results of scenarios with 30 EUR-interacting causal SNPs excluded and tagging SNPs included in the dataset**

We simulated phenotypes using the same 30 causal SNPs defined as in previous settings but restricted their effects to be interacting with EUR ancestry proportion. We applied PFstatistics to a dataset where the 30 SNPs are excluded. Again, tagging SNPs are defined as the top 15 non-causal variants with the highest absolute correlation to any of the 30 EUR-interacting causal SNPs in the sample dataset. In panels a-d, the Y-axis numbers, labels and colors denote SNP indices and their correspondence to homogeneous causal SNPs. **a.** The mean W-statistics of tagging SNPs resemble the trends of their corresponding causal SNPs. **b.** Tagging SNPs tend to be selected by PFstatistics when causal variants were absent, and the selection decisions exhibit similar overall patterns to those observed for causal SNPs. **c.** The correlations between tagging and causal SNPs are shown overall and within stratified groups. **d.** Minor allele frequencies (MAF) of tagging SNPs are displayed across stratified groups and the full sample. Groups labeled in X-axis of c and d are defined by the estimated ancestry proportion greater than 75%; individuals not meeting this threshold are categorized as admixed.

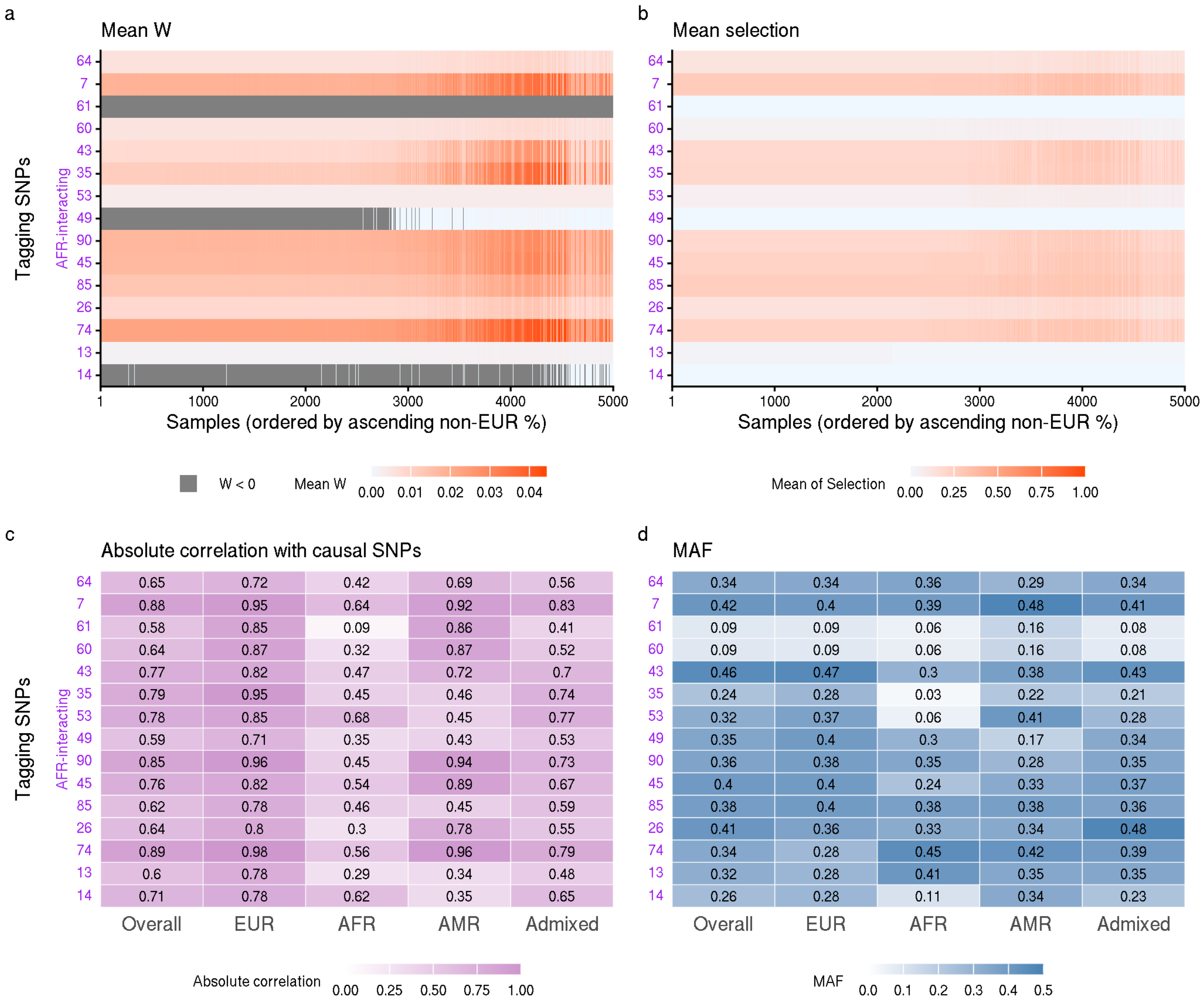


**Figure S8. Simulation results of scenarios with 30 AFR-interacting causal SNPs excluded and tagging SNPs included in the dataset**

We simulated phenotypes using the same 30 causal SNPs defined as in previous settings but restricted their effects to be interacting with the estimated AFR ancestry proportion. We applied PFstatistics to a dataset where the 30 SNPs are excluded. Again, tagging SNPs are defined as the top 15 non-causal variants with the highest absolute correlation to any of the 30 AFR-interacting causal SNPs in the sample dataset. In panels a-d, the Y-axis numbers, labels and colors denote SNP indices and their correspondence to homogeneous causal SNPs. **a.** The mean W-statistics of tagging SNPs resemble the trends of their corresponding causal SNPs. **b.** Tagging SNPs tend to be selected by PFstatistics when causal variants were absent, and the selection decisions exhibit similar overall patterns to those observed for causal SNPs. **c.** The correlations between tagging and causal SNPs are shown overall and within stratified groups. **d.** Minor allele frequencies (MAF) of tagging SNPs are displayed across stratified groups and the full sample. Groups labeled in X-axis of c and d are defined by the estimated ancestry proportion greater than 75%; individuals not meeting this threshold are categorized as admixed.

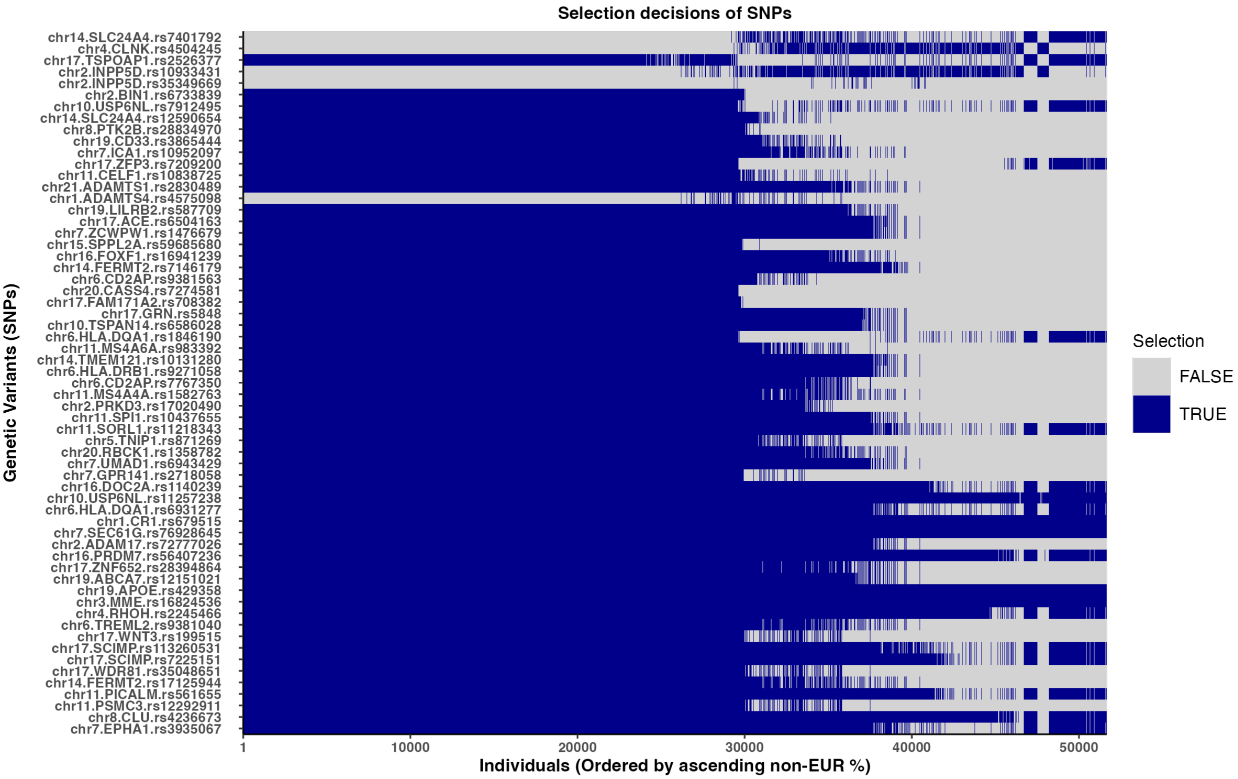


**Figure S9. Individual-level variant selection matrix generated by PFstatistics on ADSP data**

The matrix highlights heterogeneous selection patterns across individuals for SNPs selected in at least one individual in the cohort, same as SNPs present in Figure 3g. The ordering of these SNPs matches that of Figure 3g.

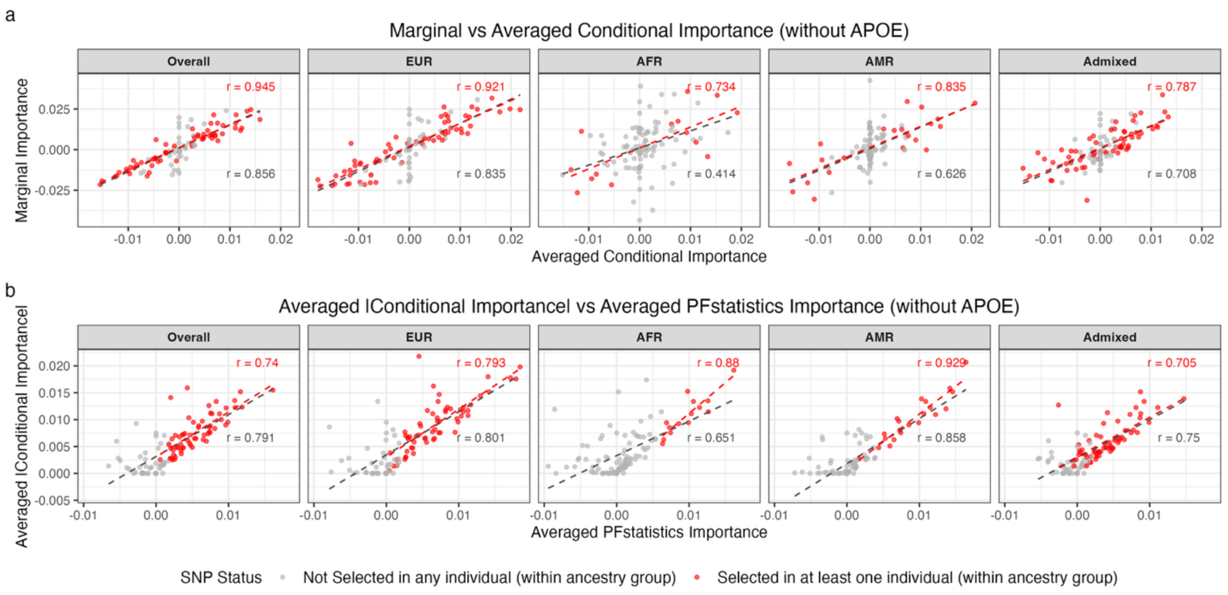


**Figure S10. Comparison between marginal, conditional, and PFstatistics importance scores across ancestry groups (without APOE)**

**a.** Scatterplots comparing marginal importance scores from ancestry-stratified Gaussian regression models with conditional importance scores derived from the Lasso interaction model, shown separately for the Overall sample and for each ancestry group (EUR, AFR, AMR, and Admixed). **b.** Scatterplots comparing in stratified groups the averaged absolute conditional importance scores with ancestry-averaged knockoff importance scores (W-statistics) produced by PFstatistics for the same SNP set. In both panels, each point represents a SNP. Red points indicate SNPs selected in at least one individual by PFstatistics within the corresponding ancestry group, whereas gray points represent SNPs not selected by PFstatistics in any individuals in that group. Red dashed lines represent the regression fitted to the selected SNPs (red points) within each ancestry group, while the gray dashed line represents the regression fitted to all SNPs. Pearson correlation coefficients (r) are reported for selected SNPs (red texts) and for all SNPs (grey texts), quantifying agreement between importance measures within each ancestry stratum.

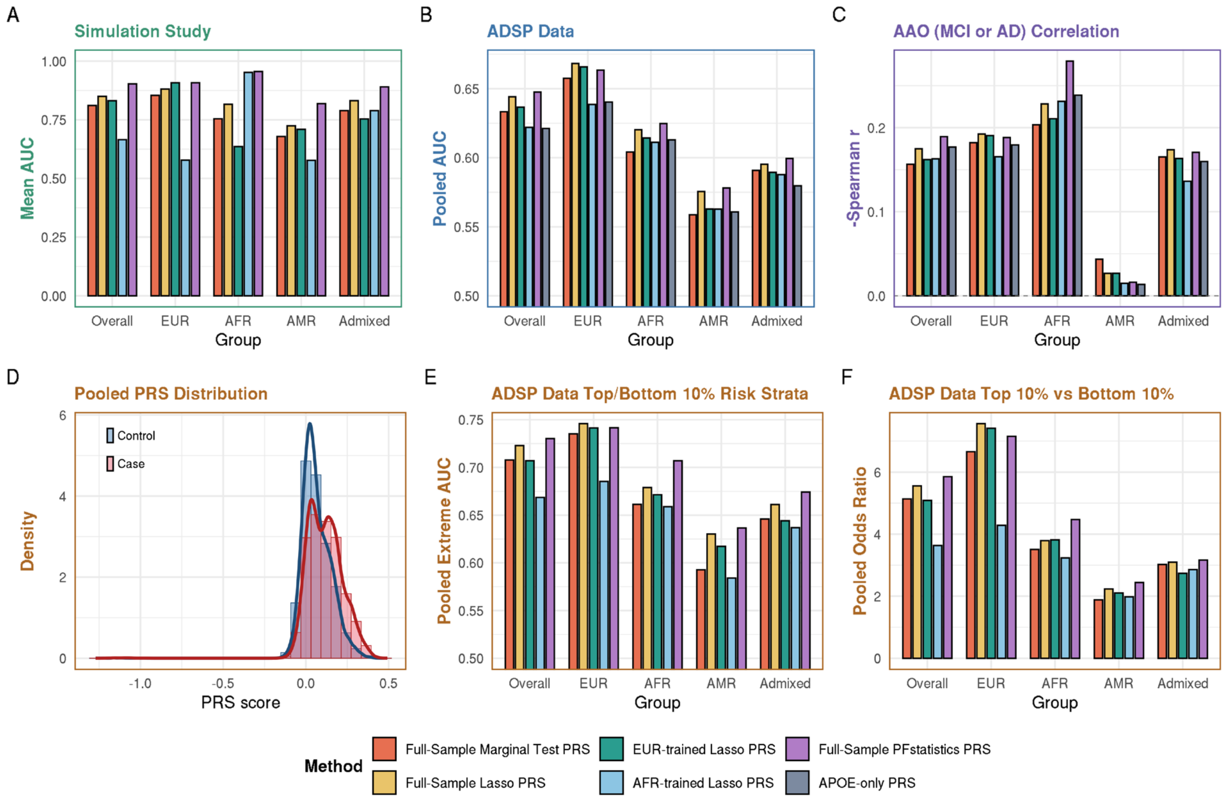


**Figure S11. Performance of PRS methods across simulation and ADSP data**

**a.** Simulation results. Bar plots show the mean AUC in left-out validation samples (n=5000). Results are stratified by overall population and by EUR, AFR, AMR, and admixed groups. **b.**

ADSP data results. Bar plots show the pooled AUC based on predictions from all five outer test folds in a five-fold outer, five-fold inner nested cross-validation framework, stratified by the overall population and by EUR, AFR, AMR, and admixed groups. **c.** Association with age at onset (AAO) among individuals with MCI or AD. Bar plots show the negative Spearman correlation between PRS and AAO, stratified by ancestry group. Larger values indicate stronger association with earlier age at onset. **d.** Distribution of pooled PRS in ADSP. Histogram and density curves show the distribution of pooled PRS scores in controls and cases. **e.** Discriminative performance in the extreme-risk strata of ADSP. Bar plots show the pooled AUC when evaluation is restricted to individuals in the top 10% and bottom 10% of the PRS distribution in each ancestry group. **f.** Enrichment of disease risk in the extreme-risk strata of ADSP. Bar plots show the pooled odds ratio comparing individuals in the top 10% versus the bottom 10% of the PRS distribution, stratified by ancestry group. Colors indicate PRS construction methods: full-sample marginal test PRS, full-sample lasso PRS, EUR-trained lasso PRS, AFR-trained lasso PRS, full-sample PFstatistics PRS, and APOE-only PRS.
